# Hematologic setpoints are a stable and patient-specific deep phenotype

**DOI:** 10.1101/2023.09.26.23296146

**Authors:** Brody H Foy, Rachel Petherbridge, Maxwell Roth, Christopher Mow, Hasmukh R Patel, Chhaya H Patel, Samantha N Ho, Evie Lam, Konrad J Karczewski, Veronica Tozzo, John M Higgins

## Abstract

The complete blood count is an important screening tool for healthy adults and is the most commonly ordered test at periodic physical exams. However, results are usually interpreted relative to one-size-fits-all reference intervals, undermining the goal of precision medicine to tailor medical care to the needs of individual patients based on their unique characteristics. Here we show that standard complete blood count indices in healthy adults have robust homeostatic setpoints that are patient-specific and stable, with the typical healthy adult’s set of 9 blood count setpoints distinguishable from 98% of others, and with these differences persisting for decades. These setpoints reflect a deep physiologic phenotype, enabling improved detection of both acquired and genetic determinants of hematologic regulation, including discovery of multiple novel loci via GWAS analyses. Patient-specific reference intervals derived from setpoints enable more accurate personalized risk assessment, and the setpoints themselves are significantly correlated with mortality risk, providing new opportunities to enhance patient-specific screening and early intervention. This study shows complete blood count setpoints are sufficiently stable and patient-specific to help realize the promise of precision medicine for healthy adults.

## Introduction

The complete blood count (CBC) is important and versatile and is ordered more frequently than any other clinical test across nearly all medical contexts^1,2^. It provides a valuable non-specific assessment of the hematologic and immunologic state of patients by measuring numbers of red blood cells (RBC), white blood cells (WBC), and platelets (PLT) per unit volume of blood, along with some basic cell population statistics. Because blood cells interrogate almost all tissues and organs with high spatial and temporal resolution, the CBC provides information on a very wide range of disease processes.

CBC indices vary dramatically among healthy adults, with the upper boundary more than twice as large as the lower in some cases (e.g., 4.5-11 x 10^3^/µl for WBC), making the one-size-fits-all reference intervals that are currently used for interpretation very wide. However, short-term intra-patient variation in CBC indices over weeks or months is significantly lower^3^. Intra-patient variation over multiple years or more has rarely been studied^4^, and it has not been directly shown whether intra-patient variation increases over decades and eventually approaches inter-patient variation. However, studies demonstrating high heritability of CBC indices^5–9^ imply that intra-patient regulatory differences between healthy patients may persist indefinitely. As the scale and scope of medical record databases advances, questions about long-term marker regulation and variation can now be investigated.

Understanding the nature of long-term variation in common diagnostic tests for healthy patients is crucial for realizing the vision of precision medicine^10,11^. Traditional medicine has always had the goal of personalizing care – but dramatic advances in diagnostic methods and data science promise much more. For healthy adults, the benefits of modern precision medicine are particularly lacking because while advanced diagnostic methods can detect many differences between healthy adults at the genetic, molecular, and cellular level, these differences have not been shown to be clinically useful other than in uncommon situations such as rare and highly penetrant Mendelian diseases. Medical care for healthy adults has not changed significantly in many years and still relies primarily on the rote application of a few one-size-fits-all population-wide screening studies.

A recent study showed that CBC results can be surprisingly stable and robust in adults, with patients who are recovering from acute inflammation reliably returning to their baseline CBC, for an extremely wide range of inflammatory stimuli including diverse trauma, ischemia, and infection.^12^ Motivating this study, we hypothesized that CBC indices in healthy individuals would be tightly regulated over long periods of times. We therefore studied CBC variation in a large cohort of healthy individuals over decades. We found that CBC indices fluctuate as expected^13,14^ but are tightly regulated around patient-specific homeostatic setpoints^15,16^. Setpoint differences between healthy individuals are sensitive to normal aging processes, chronic disease, and medical intervention, but a significant fraction of setpoint differences appears genetically determined^17,18^ and associated with novel loci. Setpoints enable clinical application of patient-specific CBC reference intervals, which show promise enhancing personalized risk assessment, and setpoints themselves are surprisingly correlated with mortality risk.

## Methods

### Patient data collection

Complete blood count (CBC) data was collected for all adult MGB outpatients over three study periods: 2002-2021, 2002-2006, 2017-2021 (Cohorts A, B, and C respectively). Patients were included if over the study period they: had >4 isolated CBCs (outpatient and >30d apart from other blood tests), had no >48h inpatient stays during the study period, and were alive at the study endpoint. Patient demographics, blood counts, procedures, medications, and diagnoses were collected using the MGB research patient data registry (RPDR), and electronic data warehouse (EDW). Diagnosis data was converted from ICD9 and ICD10 codes to disease phenotypes using PheCodes^19^. CBCs consisted of ten parameters: hematocrit (HCT), hemoglobin (HGB), mean corpuscular hemoglobin (MCH), mean corpuscular hemoglobin concentration (MCHC), mean platelet volume (MPV), platelet count (PLT), red cell count (RBC), red cell distribution width (RDW), and white cell count (WBC). MPV values were not routinely reported in the medical record before 2015 and are not reported for cohort B. Summary characteristics for cohorts A-C are given in **Table S1**. MGH reference intervals for each marker are given in **Table S2.** CBCs were run on a wide variety of hematology analyzers, reflecting changes in MGB laboratory equipment over time. Most machines typically have very low analytic variation^21^, but this may still introduce a small, fluctuating bias to marker results over time.

Each cohort was evaluated for health, with all three showing similar 1yr mortality rates (0.3%, 0.9%, 0.8% for A-C) to a similarly aged general US population (1.1% for 55-60yr olds in 2021^20^). Analysis of disease phenotypes in cohort A showed no diagnoses that would be unexpected in a general adult population of similar age – with diagnoses *pain, hypertension,* and *hyperlipidemia* being among the most prevalent (**Fig S1**).

### Setpoint calculation

The mean of a patient’s regulated healthy biological marker range (herein referred to as a ‘setpoint’) was estimated by fitting a Gaussian mixture model with up to 3 components to each patient’s set of isolated CBCs and taking the mean of the largest component. Optimal component number was chosen based on the Akaike information criteria score, and a multi-component model was selected only if the largest component proportion was greater than 70% (2 components) or 45% (3 components). The coefficient of variation (CV) was calculated from the component variance. Analytic variation will contribute to variance as well, but for modern laboratory hematology instruments is typically small (<∼1%)^21^. Inter- and intra-patient variation in blood count markers were compared to short-term intra-patient marker variation rates reported in the European Federation of Clinical Chemistry and Laboratory Medicine (EFLM) database^3^. Estimates of inter- and intra-patient marker variation in **Fig 1b-d** were calculated based on each patient’s set of isolated CBCs, without use of mixture models (to reflect overall variation, including outlier values). These results were adjusted for age-associated drift using linear regression over the entire study cohort. Rates of age-associated drift were low for all markers (**Table S3**). Unless otherwise specified, other results did not use age-adjustment, to enable unbiased comparison to reference intervals.

**Figure 1.**
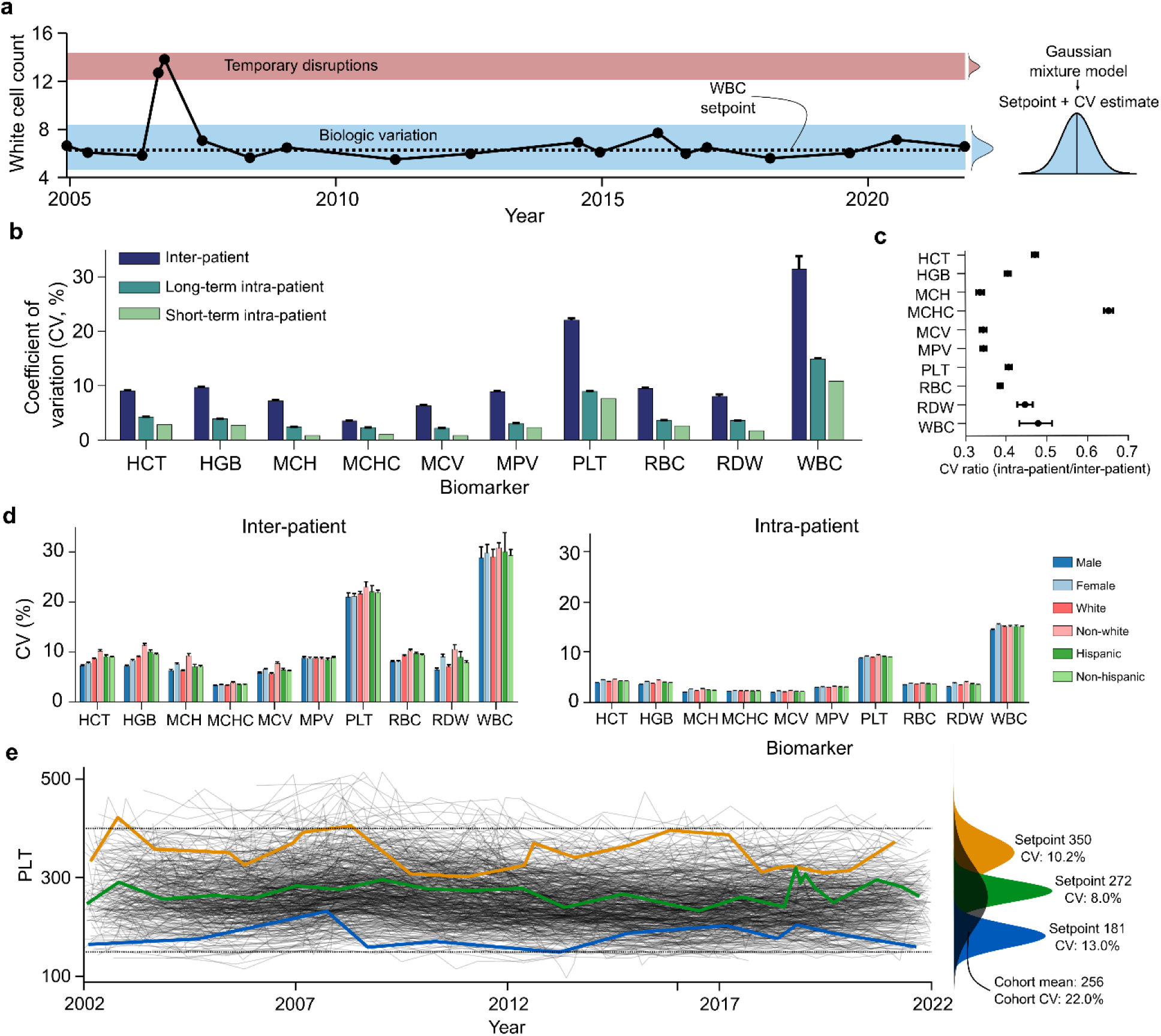
Hematologic setpoints are conserved over decades in states of health. a. A single outpatient WBC trajectory over 20yrs, showing marked stability around 6×10^3^/µL with occasional transient pathophysiologic disruptions, and a 95% confident interval (4.5-8.1) about half as wide as the adult reference interval at the study hospital (4.5-11.0). b. Inter- and intra-patient variation in outpatient marker values over 20yrs (long-term) and multiple weeks (short-term); short-term intra-patient estimates were derived from the EFLM database. c. Ratio of intra- and inter-patient marker CVs, consistently below 0.5 for most markers. D. Long-term intra-patient CV estimates by demographics – showing high consistency between groups. e. Long-term PLT trajectories for 500 randomly chosen patients, with overall distribution (black) and individual trajectories of three patients with high, moderate, and low setpoints. Error bars in b-d reflect 95% confidence intervals on the mean, calculated via bootstrapping. Stratification of patient CVs by age and over different lengths of time are given in **Fig S2-S3**. Dotted lines in e reflect the MGH reference interval. Equivalent plots to panel e for RBC and WBC are given in **Fig S6**.

### Setpoint association analyses

Associations of setpoints with other hematologic markers were analyzed using raw CBC datafiles. From Jan-2017 onwards raw datafiles from all MGH CBCs run on Sysmex XN-9000 systems were stored. 27 additional hematologic parameters were extracted from the headers of these raw data files^22^. Setpoints for each Sysmex parameter were calculated for all patients in cohorts A-C that had at least 5 isolated values available – the N of this varied by parameter as for any individual CBC only a subset of the Sysmex parameters is typically reported into the file header. Data availability and summaries for these parameters are given in **Table S4.**

Associations of setpoints with non-hematologic markers were analyzed via a matched prospective study. Healthy patients in cohort C attending MGB system outpatient visits were identified and matched to demographically similar patients with a significant difference in their HCT, PLT, RDW or WBC setpoint.

Using excess clinical specimens, a series of general inflammatory, renal, liver and serum lab markers were collected. Differences between matched patient pairs were analyzed using 2-sided t-tests and hierarchical clustering. Full details of enrolment criteria can be found in the **Supplementary Methods**. Characteristics of the prospective cohorts are given in **Table S5**.

### Physiologic shifts

We tested whether physiologic changes and pathophysiologic states could alter setpoints by analyzing CBCs of patients in cohort A for whom menopause was noted or who were diagnosed with hypothyroidism or liver disease (any diagnosis of hepatitis, cirrhosis, or chronic liver disease), or who had a splenectomy procedure. These four cohorts were each hypothesized to exhibit setpoint alterations based on the associated physiologic or pathophysiologic change. For each diagnosis, patients were included if they had at least 5 isolated CBCs pre-event (study start to 1yr pre-) and post-event (1yr post to study end). We also investigated setpoint changes pre- and post-pregnancy by considering all patients across the four cohorts in this study who had a delivery recorded in a local obstetrics database (containing 40,000+ MGH-associated deliveries)^23^. In this cohort, setpoint changes were analyzed using all adult isolated CBCs at least 2yrs pre- or 2yrs post the date of delivery, excluding patients with fewer than 5 isolated CBCs in each period. Setpoints were calculated pre- and post-event after correction for age-associated drift in lab values (via linear regression, **Table S3**). Characteristics of these cohorts are given in **Table S6**.

### Heritability analysis

Setpoint heritability was estimated using patient relationship data in the electronic health record (EHR), similar to prior reports^24^. All patients in cohorts A-C with first-degree familial (parent-child or sibling) or partner (spouse or life partner) relations also in one of the cohorts were retained (N: 439, 440 pairs respectively). Familial relationships were assumed biologic unless otherwise noted (e.g., stepfather, etc.). Heritability was estimated as 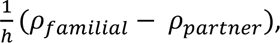 where *ρ_familial_* and *ρ_partner_* are the correlations between familial and partner setpoints, and ℎ is the genetic strength (0.5 for first-degree relatives), with *ρ_partner_* acting as a control for environmental effects. All setpoints were age- and sex-corrected via linear regression prior to correlation estimation. Heritability estimates were compared to literature estimates from five studies: 2 twin studies^5,6^, 1 pedigree study^7^, 1 multi-generation study^8^, and 1 large-scale EHR study^9^. Estimates were also calculated using a randomly chosen isolated CBC from each patient. Corresponding heritability estimates are given in **Table S7**.

### Genome-wide association studies

To assess setpoint-genome associations, data from the MGB Biobank – a biorepository of genotyped samples from consented MGB patients – was used. All Biobank patients who had genotype data, at least 5 isolated CBCs, and who were alive as of Apr-01-2023 were included (N: 32,093). Patient genotyping was performed by the MGB Biobank, using the Illumina Multi-Ethnic Genotyping (MEGA) and Expanded MEGA (MEGA^EX^) array covering 1,416,020 and 1,741,376 SNPs respectively. Results were imputed by the MGB Biobank team using the Minimac3^25^, on the Michigan Imputation Server, with the Haplotype Reference Consortium (r1.1 2016) panel. Samples with high SNP missingness (first pass filter 0.2, second pass filter 0.02), high heterozygosity, sex discrepancy, or high relatedness (kinship coefficient >0.2), and those of non-European ancestry (identified by comparison of multidimensional scaling plots to the 1000 Genomes reference panel^26^) were excluded (due to insufficient sample size for analysis in non-European subjects). SNPs with low minor allele frequency (<0.05), poor imputation (R^2^ < 0.3), or outside Hardy-Weinberg equilibrium (p<1e-10) were excluded. GWAS analysis was performed using a linear model with age, sex, and 10 genetic principal components as covariates. Setpoints were calculated using the same procedure as described in prior sections. Analysis was also performed using a randomly chosen isolated CBC from each patient. Loci were identified using the plink *clump* feature with significance thresholds of 5e-8 and 1 for primary and secondary SNPs, a linkage disequilibrium threshold of R^2^ > 0.2, and a clumping region of 250kbp. A locus was deemed novel if there were no reported significant associations to any of the 9 CBC markers for any SNP within 250kbp of the locus’ primary SNP, as determined by queries to the GWAS catalog^27^. All novel loci were subsequently manually reviewed to confirm novelty. SNP heritability estimates were derived from the GWAS summary statistics using the *sum-hers* function in LDAK^28^ version 5.2, with the LDAK-Thin tagging file (as supplied on the LDAK website^29^) and excluding any predictors which explained more than 1% of phenotype variation. All other analysis was implemented using *bcftools*^30^*, plink* and plink2^31^, with a significance threshold of 5e-8 unless otherwise specified. Significant hits and loci from each GWAS are provided in **Tables S8-S10**, and full GWAS results can be accessed via GWAS catalog (accession numbers are given in the data and code availability statement). Summary characteristics of the MGB Biobank cohort are given in **Table S1**.

### Statistical analysis

Statistical significance of data was calculated using t-tests for continuous variables, and chi-square for categorical variables. Survival analysis was performed using Kaplan-Meier curves, with statistical significance calculated using log-rank tests. For time-to-event analysis, patients were censored at the date of last data collection (Jan-01-2023 unless otherwise noted). Thresholds for significance for all analyses were set at p = 0.05 unless otherwise noted. All non-genomic data analysis was performed using MATLAB 2021b.

## Results

### Complete blood count indices are tightly regulated in healthy adults over decades

We studied intra-patient variation in CBC indices over a 20year period (**Fig 1a**). Standard CBC reference intervals are based on inter-patient CVs of CBC indices, which ranged from 10-30% in our cohort (**Fig 1b**). Intra-patient CVs were much lower over the 20year period, only 30-70% as large as the equivalent inter-patient CV (**Fig 1c**). Intra-patient CVs over 20 years remained closer to previously reported intra-patient CVs determined over much shorter periods of a few weeks or months^3^. The magnitude of the CV did not vary with gender, demographics, or age (**Fig 1d, Fig S2-S3**), consistent with the hypothesis that this degree of regulation is a generic feature of normal physiology. The autocorrelation function for each CBC index decayed more slowly than a random walk process, consistent with an active regulatory process (**Fig S4**). The CVs were also stable for a wide range of values for each setpoint (**Fig S5**), consistent with the hypothesis that the regulatory process was equally efficient for a range of setpoints. Thus, each patient’s CBC indices appear to be regulated to stay within a sub-interval of the inter-patient range for decades, and these hematologic setpoints represent a well-defined state of health for each patient (**Fig 1e, Fig S6**).

### Hematologic setpoints vary significantly among healthy patients

The three patients in **Fig 1e** can be distinguished for most of the 20year period based on just their PLT count. Considering all 9 setpoints (excluding MPV due to more limited availability), for the average patient only 2% of the cohort had all setpoints within that patient’s setpoint intervals (setpoint ± 2CV). Thus, a healthy patient’s set of hematologic setpoints is highly specific and identifies a personalized state of health that is typically distinct from that of more than 98% of other healthy individuals.

This high degree of patient-specificity reflects modest intra-patient correlations between setpoints and other non-routine hematologic parameters (**Fig 2a**). There is, as expected, strong correlation for setpoints with co-dependent definitions (e.g., HCT, HGB, RBC, or WBC, ALYMPH, ANEUT), physiologic trade-offs (e.g., RBC vs MCH and MCV; PLT vs MPV), and between MCH and MCV, given MCHC is tightly regulated across all vertebrates^31^. However, RDW was inversely correlated with other red cell setpoints, suggesting increased cell size and production are associated with reduced size variability. RDW was also correlated with immature reticulocyte fraction, and fragmented and nucleated red cell counts, suggesting chronic RDW elevation may reflect sustained dysregulated production or elevated schistocytosis^32^, potentially explaining part of the well-known RDW-mortality association^33^. MPV was positively associated with immature platelet fraction and platelet distribution width, possibly reflecting a negative association with PLT age^34^. WBC was associated with relative lymphocyte counts, suggestive of some chronic inflammation in patients with elevated setpoints. These results highlight that CBC setpoints may often reflect sustained differences in underlying cell production, clearance, and regulation features, whether acquired or genetic.

**Figure 2.**
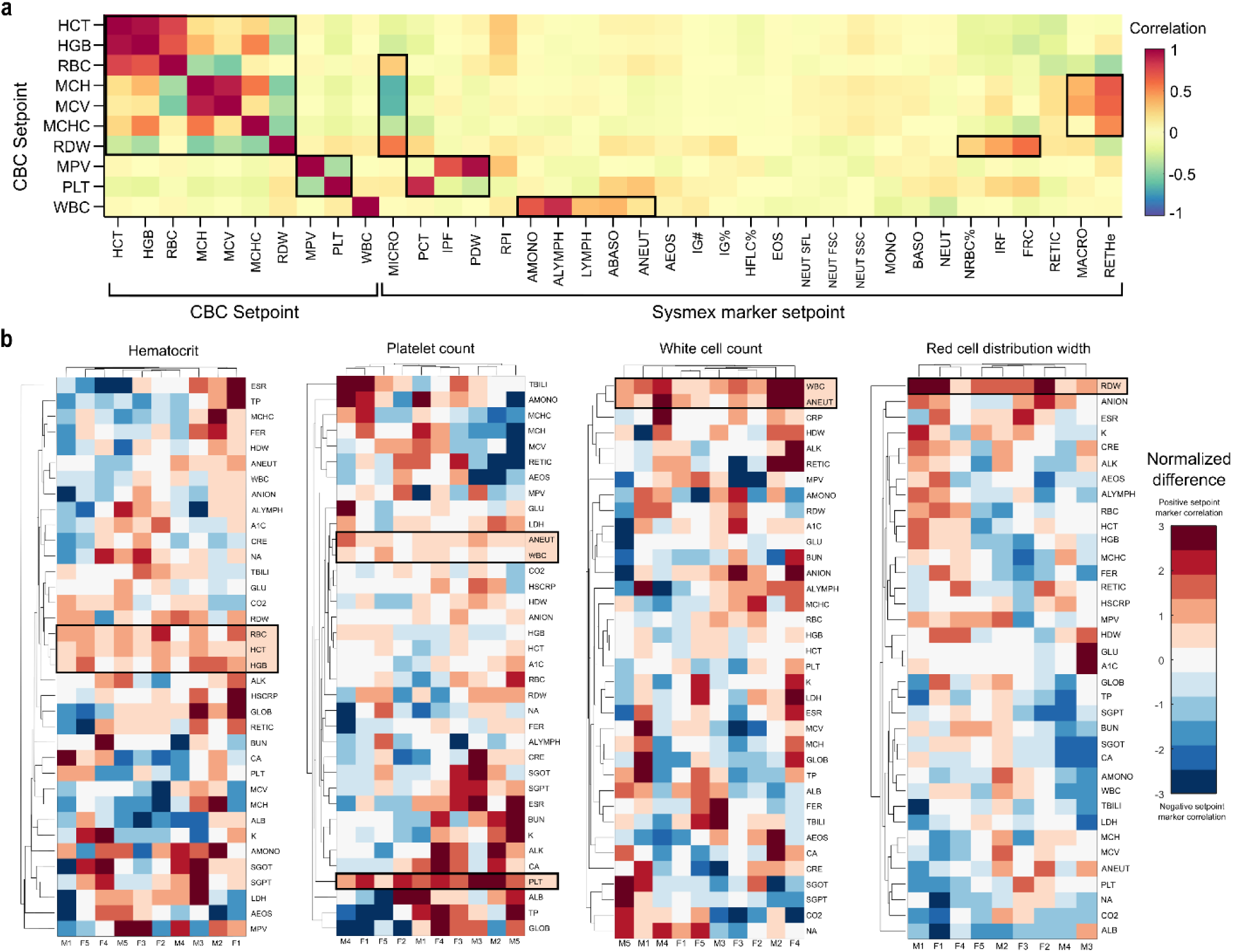
Physiologic associations and determinants of setpoints. a. Correlations between setpoints and broader hematologic parameters from the Sysmex XN-9000. b. Comparison of lab marker differences across 10 matched patient pairs with setpoint differences. Markers in both a and b were ordered by hierarchical clustering. Highlighted values in b reflect those with p<0.05 significance (2-sided t-test). Summary characteristics for the Sysmex markers and four prospective cohorts are given in **Table S4-S5**. Note that RDW results in b are limited to 9 patient pairs due to resource constraints (see Supplementary Methods for further details).

### Acquired origins of setpoint variation

The heterogeneity of hematologic setpoints across healthy adults raises the question of the origin of these inter-patient differences, and the extent to which acquired or genetic factors may be involved. Differences in disease history or exposures could potentially alter hematologic setpoints. For instance, chronic disease is often associated with low-level inflammation which might raise the WBC setpoint, along with other inflammatory markers. To test this hypothesis, we prospectively measured a panel of routine lab tests in a set of matched patients where one patient had a low hematologic setpoint (HCT, PLT, WBC, or RDW) and the other high. (See Supplemental Methods for more detail.) No associations were seen between setpoints and common metabolic, inflammatory, renal, or liver markers. This small study does not exclude the possibility of an environmental contribution to setpoint differences but suggests a large contribution of this type is less likely. Consistent with prior studies^35^, CBC indices showed small age-related changes with most below 1% CV per 10 years (**Table S3**), though setpoint CVs were not significantly age-dependent (**Fig S2**). Additionally, some normal physiologic changes like pregnancy and menopause were associated with setpoint shifts (**Fig 3a-f**). Menopause was associated with a HCT setpoint increase, consistent with reduced blood loss and improved iron status^36^. Pregnancy was associated with decreased MCHC setpoints post-partum (**Fig 3f**), a novel finding which suggests the transient increased erythropoietic demand of pregnancy may lead to persistent changes in erythropoiesis.^37^ Chronic disease states like hypothyroidism and hepatitis, and medical interventions such as splenectomy were also associated with changes in hematologic setpoints that are consistent with expectation and prior observations (elevated MCV^38^, reduced PLT^39^ and increased RDW^40^ respectively). In each of these cases, the use of a setpoint instead of a single CBC marker to detect a change yielded similar effect size estimates, but dramatically increased statistical significance (**Fig 3g**), suggesting setpoints enable greater sensitivity and precision for detecting and estimating changes in patients’ states of health. These results demonstrate that setpoints are modifiable, but the magnitudes of change are modest, leaving the majority of setpoint variability in the healthy adult population unexplained.

**Figure 3.**
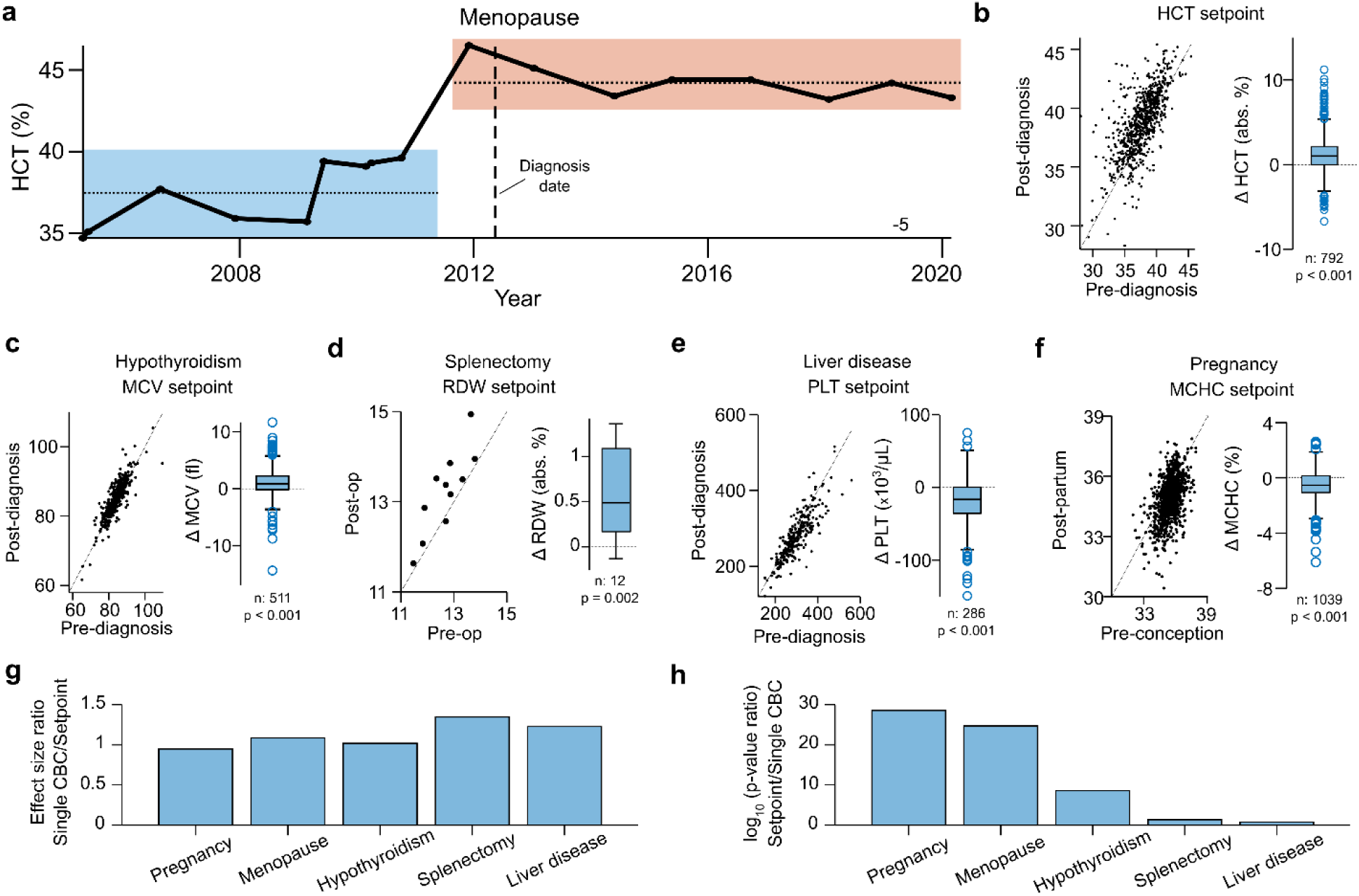
Setpoint shifts across various pathophysiologic settings. a. Long-term HCT trajectory in a patient pre- and post-menopause, illustrating a clear shift in HCT setpoint. b-f. Shifts in patient setpoints pre- and post-pathophysiologic events: c. hypothyroidism, d. splenectomy, e. liver disease, f. pregnancy. g. Ratio of effect size (mean marker change) estimates when using setpoints compared to a randomly chosen single isolated CBCs. h. Ratio of p-values from a t-test of effect sizes using setpoints or isolated CBCs, on a log_10_ scale. Summary characteristics of cohorts in b-f are given in **Table S6**. Lines in b-f reflect unity.

### Genetics origins of setpoint variation

Given the limited role of acquired factors, we investigated the genetic basis of setpoint differences by performing heritability and GWAS analysis. We first used EHR-derived familial relationships within cohorts A-C and found that setpoints were highly correlated between first-degree relatives but not partners (a proxy for environmental effects) (**Fig 4a-b, Fig S7-S8)**. Except for MCHC, all CBC setpoints showed strong heritability (**Fig 4c**). Estimating heritability from EHR-derived relationship data can be noisy, but estimates using setpoints in these cohorts were consistent with mean results published previously^5–9^ (**Table S7**), and closer to the literature mean than single-CBC estimates except for RDW (where both estimates were extremely close to the mean).

**Figure 4.**
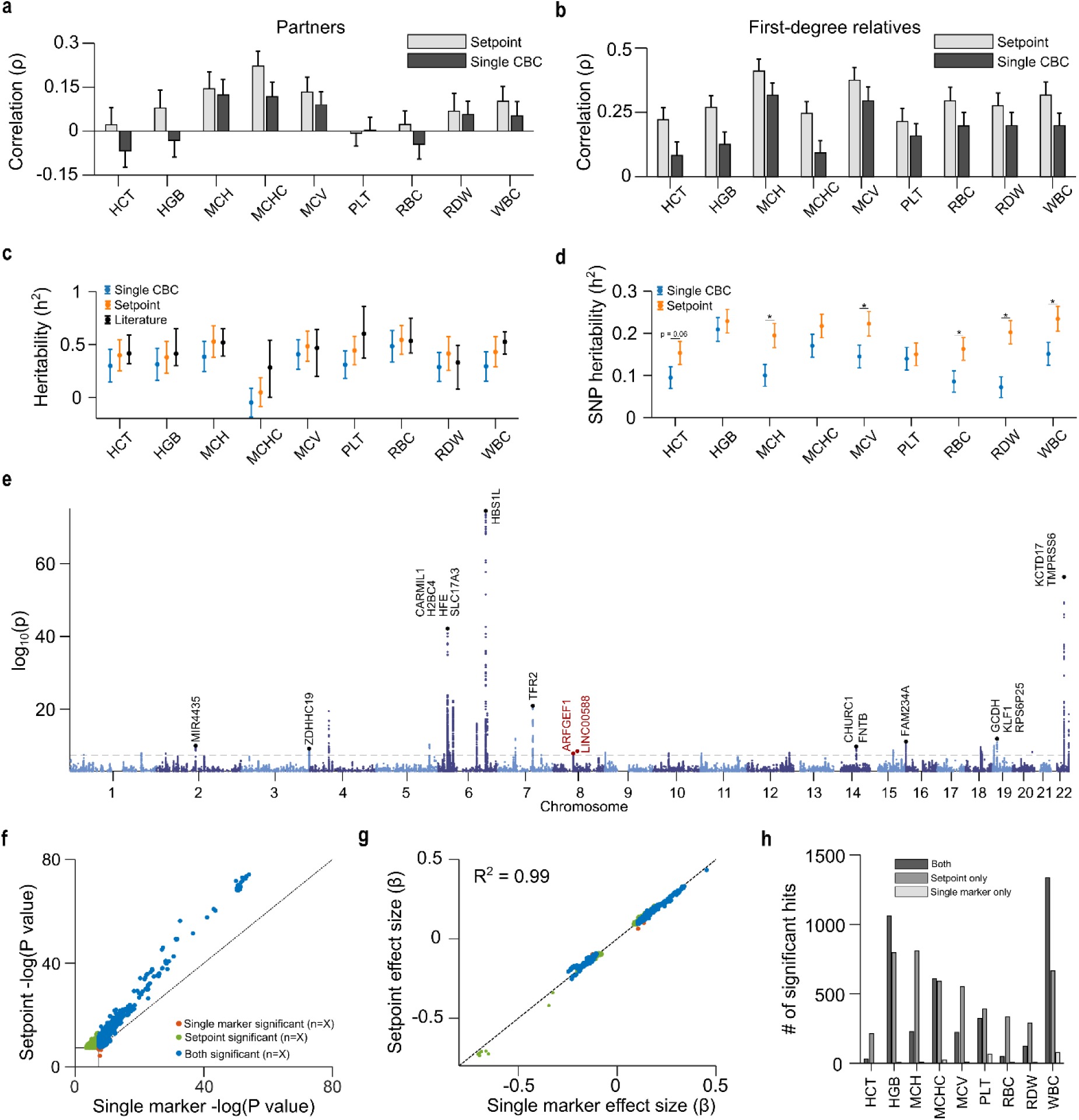
Setpoints reflect a deep phenotype, heightening mechanistic discovery. a-b. Setpoint and single CBC correlations between romantic partners (a) and first-degree relatives (b). c. Heritability estimates derived from setpoints and single CBCs compared to literature values. d. SNP-heritability estimates from a cohort of 25,254 MGB patients, using setpoints and single CBCs. e. Manhattan plot for a GWAS of HGB setpoints in a cohort of ∼25,000 MGB patients. f-g. Comparison of p-values (f) and effect sizes (g) in GWAS using HGB setpoints and single outpatient values. h. Yield increases in significant hits using setpoints comparative to single CBC markers. Dashed lines in f-g correspond to unity. Annotations in panel e correspond to nearby genes for highly significant association loci, with red annotations corresponding to novel loci. Literature heritability estimates are given in **Table S7**. Raw data plots for a are given in **Fig S7-S8**. Equivalent plots of panels e-g for other setpoints are given in **Fig S9-S10, S16-S17**. Quintile-quintile plots for each setpoint GWAS are given in **Fig S18**. A full list of GWAS hits, association loci and gene contexts is given in **Tables S8-S10**.

We also performed a GWAS using both setpoints and single CBC measurements. SNP-based heritability estimates were consistently higher when using setpoints (**Fig 4d**), potentially explaining part of the well-known missing heritability problem for these indices^41^. GWAS of the 9 CBC setpoints identified 397 associated loci. Eight loci (lead SNPs: rs60528951, rs2047265, rs12522573, rs6997857, rs10021975, rs10043270, rs117912622, rs869243453) appeared to be novel after comparison to the GWAS catalog^27^ and to results derived from much larger studies that relied on 1-2 CBC measurements^17,18^ (**Fig 4e**, **Table S8-S10**). Setpoint GWAS produced comparable effect size estimates to single marker GWAS but with increased significance (**Fig 4f-g, Fig S9-S10**), leading to an average 3.6-fold increase in significant SNPs (**Fig 4h**). Setpoints also provided improved discovery over 2, 4, and 8-CBC averages, suggesting that identification of the underlying biologic distribution and systematic exclusion of outlier values is worthwhile (**Fig S11**).

### Setpoints provide population-wide and personalized information on clinical risk

Hematologic setpoints are thus stable for decades and patient-specific, with differences between healthy patients likely reflecting contributions from genetics, normal aging, and possibly exposure history. We first investigated whether these different patient-specific healthy states were clinically meaningful. When limited to ‘normal’ (within reference interval) values, setpoints showed consistent and significant monotonic associations with mortality (**Fig 5a-b**, **Fig S12**), even after controlling for age and sex (**Fig 5c**), and under replication in an independent cohort (**Fig S13**). After controlling for age, sex, and setpoint value, increased setpoint variability was also associated with higher mortality risk (**Fig 5d**) for all markers except MCHC (which had very low variability for nearly all patients). Most risk associations between setpoints and mortality had the expected directionality (e.g., low PLT and high WBC both associated with higher risk), but higher MCV was unexpectedly associated with elevated risk.

**Figure 5.**
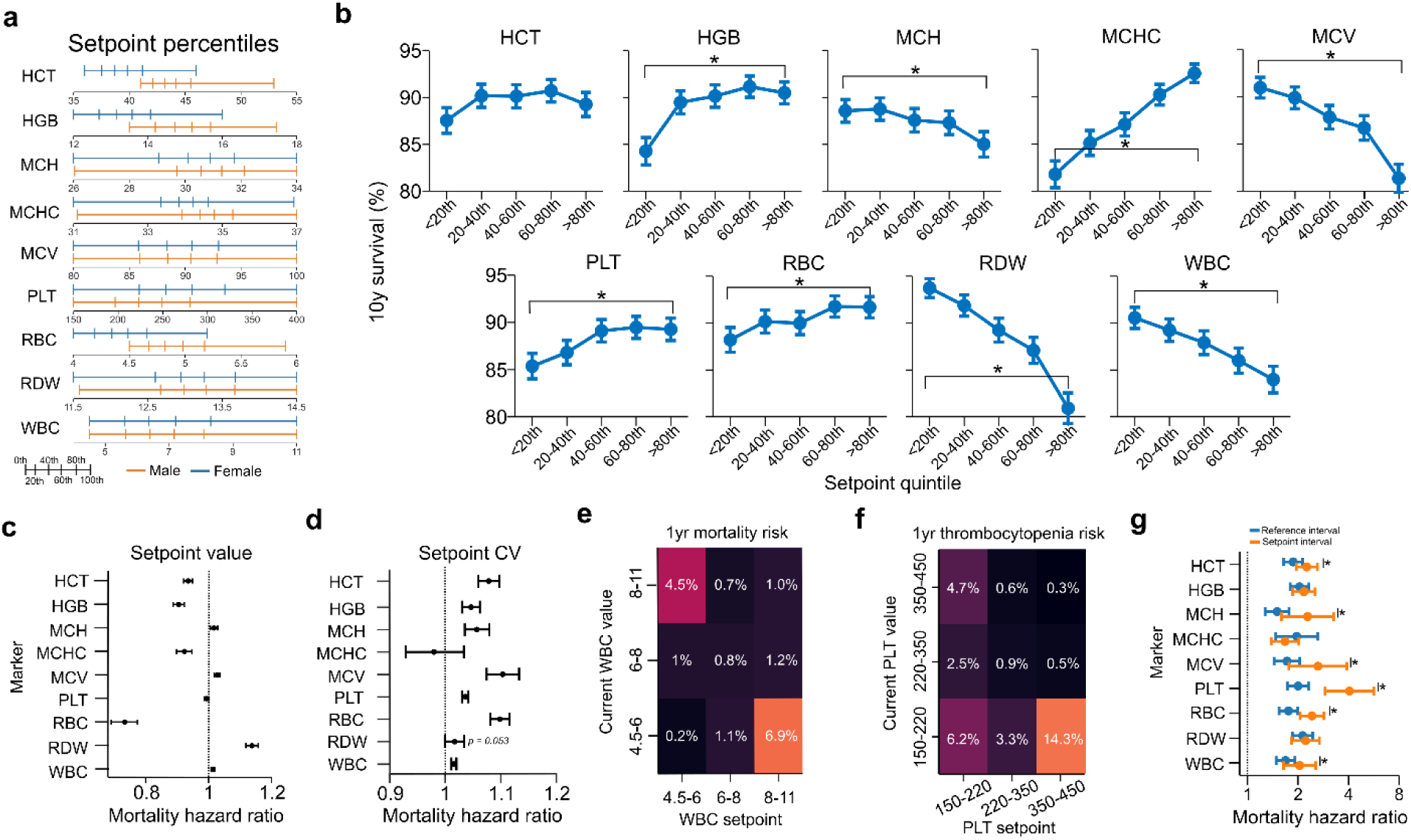
Hematologic setpoints capture significant aspects of patient mortality risk. a. Setpoint distributions across cohort stratified by sex. b. 10yr mortality stratified by setpoint quintiles, excluding patients with abnormal setpoint values. c-d. Age- and sex-corrected 10y mortality hazard ratios by setpoint value (c) and coefficient of variation (d). e. Likelihood of mortality within 1yr given a patient’s current WBC count and setpoint. f. Likelihood of future thrombocytopenia (PLT < 150×10^3^/µL) given current PLT count and setpoint. g. 10yr age, sex, and setpoint-corrected mortality hazard ratio if the current marker is outside the MGH reference interval (blue) or more than ±2std away from the setpoint. Results in c-d exclude setpoints outside the MGH reference interval. Results in d were corrected for the associated setpoint value. Results in g use the mean intra-patient standard deviations calculated in Fig 1e. Equivalent results to panels e-f are provided for a range of markers in **Fig S14-S15**. Results for b over different time periods are given in **Fig S12**.

Setpoints may also allow for improved interpretation of new test results. When comparing WBC setpoints in cohort B (calculated between 2002-2006) to a patient’s worst outpatient lab value in 2007, 1yr mortality was greatest if the patient had a high setpoint and low current value (6.9%) or vice versa (4.5%) (**Fig 5e**), with similar results seen for multiple other markers (**Fig S14**). Similarly, when stratified by current PLT value and setpoint, future risk of thrombocytopenia (PLT<150×10^3^/µL) was highest for patients with a high setpoint and low current value (**Fig 5f**) (with similar seen for other markers, **Fig S15**). These non-linear interactions between setpoint and current marker values suggest that in many cases deviation from the setpoint may be a stronger prognostic signal than a stably high or low hematologic state.

We therefore investigated whether a personalized setpoint reference interval (setpoint ± 2CV, equivalent to a 95% confidence interval) may provide more prognostic value than a standard reference interval. For all markers except MCHC, the setpoint interval provided a higher point estimate of mortality risk than the generic reference interval, with most differences being statistically significant (**Fig 5g**). This suggests that setpoints may allow for more personalized benchmarking of patients in clinical contexts.

## Discussion

We find that CBC indices in healthy adults fluctuate narrowly around setpoints for decades. 95% confidence intervals derived from setpoints are typically less than half the width of 95% confidence intervals for the population. Patient-specific intervals are highly personalized, with the set of each patient’s intervals excluding at least one setpoint from more than 98% of other healthy adults in our study cohort.

Biologic variation of hematologic markers has been well studied over shorter durations of a few weeks or months^3,4,42^, but few studies have assessed intra-patient variation in large cohorts over multiple years or more^4^. CBC indices are known to exhibit age-associated drift^35,43^, and our study shows both that patient-specific setpoints can be adjusted for this drift (**Fig 1**), and that the magnitude of age-associated drift is modest compared to inter-patient heterogeneity in setpoints (**Table S3**). Estimates of intra-patient setpoint variation were uniform across patient age, sex, and self-reported race or ethnicity (**Fig 1b-d, Fig S3-S4**), suggesting hematologic regulatory systems are fundamental to healthy physiology, despite differences in burden of chronic disease that might persistently alter hematologic state^44^. This stability and patient-specificity of CBC setpoints supports their use in specialized intra-individual monitoring contexts like the athlete biological passport used to detect doping in professional athletics^45^. Harris et al., suggested that an index of individuality (the ratio of intra- to inter-patient marker CV) below 0.6, is evidence that generic reference intervals may be insufficient^46^. Results in **Fig 1c** show that most hematologic setpoints have a long-term index of individuality well below this threshold, highlighting inherent limitations of the currently used generic reference intervals.

The stability and specificity of setpoints provides opportunities for the personalized risk assessment and treatment of healthy adults envisioned by precision medicine^10,11^. Our analysis shows that setpoint-derived patient-specific reference intervals provide more accurate mortality risk prediction than population-wide reference intervals (**Fig 5g**). These results motivate future studies to determine if clinical outcomes may be improved by substituting patient-specific CBC reference intervals into clinical management guidelines that currently rely on population-wide CBC reference intervals. Other studies have shown potential value of multi-dimensional hematologic reference intervals^47^ – motivating future study of multivariate setpoint intervals.

Setpoints levels themselves stratify patients by mortality risk (**Fig 5a-c**). The monotonic increase in mortality risk seen for quintiles of several setpoints is unexpected. Few physicians would expect that a difference in MCHC of 33 mg/dL vs 34 mg/dL would be associated with a 40% relative increase in 10yr mortality (14.6% for MCHC 33.5-34.5% vs 10.5% for MCHC 34.5-35.5%; N = 1698, 4504; mean(std) age: 57.8(16.1), 57.5(15.6), P=5e-6). These results suggest that CBC setpoints are continuous measures of patient state and that no population-wide threshold can optimally capture patient risk. Precision medical care of healthy adults^9–11^ would thus integrate setpoint-based reference intervals as well as the setpoints themselves. These results motivate further study to determine which diseases are responsible for this increased mortality, and whether it would be effective to introduce additional screening measures for patients that would be triggered by their setpoint values. Further understanding of setpoint-associated differential mortality risk may also indicate whether interventions intended to modify setpoints, as is shown to be possible (**Fig 3**), might be helpful.

CBC setpoints show high heritability, consistent with what has been shown for CBC indices previously, and GWAS efficiently identifies hundreds of associated loci. Greater understanding of the genetic architecture of setpoints (and analysis in non-European ancestry populations) might eventually enable inference of an individual’s setpoints from genotype data if serial CBCs were not available. The loci and SNPs associated with setpoints may also provide clues about the diseases responsible for differential mortality risk associated with setpoints (**Fig 5a-c**). Given our modest cohort size (comparative to prior GWAS studies of hematologic markers^17,18^), the identification of 8 novel loci highlights the strength of the physiologic signal provided by hematologic setpoints as compared to individual measurements (**Fig S16**). More broadly, these results provide a clear example of the value of deep phenotyping for GWAS, aligning with recent studies in this area^48–50^.

While CBC indices are tightly regulated in healthy adults, they are far from constant. The correlation between intra-patient variance and mortality risk may reflect allostasis or different allostatic loads across patients^15^. The identity of these setpoints and their correlation structure (**Fig 2a**) also provide opportunities for future investigation of the underlying mechanisms driving inter-patient setpoint differences, for instance determining whether variation is greater between patients in terms of rates of production, lifespan of cells and platelets in the circulation, or the processes that govern clearance. Other work has suggested that hormonal factors may influence hematologic setpoints^43^, a clear area for future study.

Here we have shown that in healthy adults hematologic setpoints are stable and tightly regulated over decades. These setpoints reflect deep phenotypes of hematologic regulation, which can heighten discovery in mechanistic and associational studies. Setpoints are novel and generic markers of patient risk, and also provide opportunity for improved care through patient-specific benchmarking. Hematologic setpoints provide opportunity to realize the goal of precision medicine, while utilizing data that is already routinely measured worldwide.

## Data Availability

Due to restrictions on sharing protected health information, individual patient data has not been shared. Supplemental methods, figures and tables are given in Supplemental File 1. GWAS summary data has been uploaded to the GWAS catalog and is available at accession number (to be inserted in later pre-print version). Data for significant hits and loci is given in Supplemental File 2. Code for calculating of setpoints is included in Supplemental File 3.

## Acknowledgements

We thank the Mass General Brigham Biobank for providing access to genomic and health information data, and the MGB Electronic Data Warehouse and Research Patient Data Repository groups for facilitating access to electronic health records. We thank Rahul Gupta and Jon Stefely for helpful discussions. We thank Kent Lewandrowski and the MGH core clinical laboratory for enabling prospective laboratory testing.

## Ethics

The study protocol was approved by the local institutional review board at Massachusetts General Hospital

## Funding + conflicts of interest

JMH reports funding from the National Institutes of Health (grant IDs: R01HD104756; R01DK123330). All authors report no conflicts of interest.

## Author contributions

JMH and BHF conceived the project and its design. Data collection was performed by BHF, RP, VT, MR, HRP, CHP, SNH and JN. Data analysis was performed by BHF, with input from all authors. All authors contributed to interpretation of results, writing and editing of the manuscript.

## Data and code availability

Due to restrictions on sharing protected health information, individual patient data has not been shared. Supplemental methods, figures and tables are given in **Supplemental File 1**. GWAS summary data has been uploaded to the GWAS catalog and is available under accession numbers: GCST90292591 (HCT), GCST90292592 (HGB), GCST90292593 (MCH), GCST90292594 (MCHC), GCST90292595 (MCV), GCST90292596 (PLT), GCST90292597 (RBC), GCST90292598 (RDW), and GCST90292599 (WBC). Data for significant hits and loci is given in **Supplemental File 2**. Code for calculating of setpoints is included in Supplemental File 3.

## Notes

### Competing Interest Statement

The authors have declared no competing interest.

### Author Declarations

Institutional review board of Massachusetts General Hospital gave ethical approval for this work

### Summary of Updates

Updated the manuscript to include GWAS catalog accession numbers for the GWAS results.

