## Supplemental File 1 for "Hematologic setpoints are a stable and patient-specific deep phenotype"

Contents

- Supplemental methods
- Supplemental figures S1-S17
- Supplemental tables S1-S7

Note: Due to size, **Table** **S8, S9** and **S10** are included in excel file **Supplemental File 2**.

Supplemental methods

**Prospective study**
Hematologic setpoint differences between healthy patients may arise as a result of both congenital and acquired contributions. If CBC setpoint differences arise to a significant degree from environmental exposures or medical history, we would hypothesize that setpoint differences might correlate significantly with levels of other routinely measured laboratory tests. For instance, if subclinical chronic inflammation is an important determinant of the WBC setpoint in healthy patients, then we would hypothesize that patients with higher WBC setpoints would be more likely to have higher levels of non-specific inflammatory markers like CRP, ESR, or LDH. We therefore performed a small prospective study to test the hypothesis that CBC setpoints for healthy individual are statistically significantly correlated with levels of other common lab tests. This study was performed by constructing four cohorts of patients, each consisting of 10 demographically matched patient pairs with significant differences in one of their HCT, PLT, RDW, or WBC setpoints. 10 patients per arm was chosen primarily due to financial and time constraints of the study, and powered detection of marker differences with standard deviation up to 80% of the mean difference.

For each setpoint, each day between Jan-01-2023 and May-30-2023, all members of cohort C who had outpatient CBC orders at MGH were identified. Patients were excluded if any of the CBC results were outside the MGH reference interval, if they’d had other bloodwork within the last 30days, or if their visit-associated bloodwork involved any non-routine test orders. Routine tests were the WBC differential, hemoglobin A1C, metabolic panel, glucose, lipid panels, liver function panels, vitamins B12 or D, magnesium, phosphorous, urinalysis, or TSH; additional tests were occasionally judged as routine by chart review of the patient). Following medical chart review, patients were additionally excluded if they: had any history of hematologic malignancy; had current or prior history of cancer with less than 5yrs remission; had a current diagnosis of a hematologic disorder (anemia, hemochromatosis, etc.); were taking an anti-coagulant (heparin, etc.); or had any comorbidity that was judged to potentially alter their hematologic state or reflect severely diminished health (e.g., pregnancy, positive HIV status, chronic obstructive pulmonary disorder).

For each setpoint cohort, the first 5 males and females were enrolled. The remaining 10 patients were only enrolled if they matched to one of the initial 10 patients on sex, age (<5yr), and HCT (<2%), PLT (<70x10^3^/µL), RDW (<1%) and WBC (<1.5x10^3^/µL) setpoints, excluding the setpoint of interest. For the setpoint of interest, matches were required to have a high value if their match had a low value and vice versa, with high and low cut-offs as the median values for cohort C: HCT: 43%/39.7% (male/female), PLT: 216/256x10^3^/µL, RDW: 13.1%/13.2%, and WBC: 6.6/6.6x10^3^/µL. Patients were only allowed to appear in one of the four cohorts, and could only be matched to one other patient.

For all enrolled patients, prospective analysis was performed using excess clinical specimens from their outpatient blood draw, by ordering the following tests: Basic metabolic panel, WBC differential, C-reactive protein, erythrocyte sedimentation rate, ferritin, lactate dehydrogenase, reticulocyte count, liver function test panel. Any tests that were ordered as part of patient care were not repeated. Patients were retroactively excluded if any of the tests could not be run due to insufficient blood volume, or lack of appropriate tube type. Differences across the cohorts were analyzed using 2-sided t-tests, and hierarchical clustering.

Due to resource constraints, the prospective study arm was closed on May-30-2023. At this time point 79 of the intended 80 patients had been enrolled, while no valid match had been found for 1 remaining male patient in the RDW sub-cohort after 93 days of searching. As such, results for RDW present 18 patients (9 pairs) instead of 20.

A summary of demographics, laboratory test values, and clinical characteristics of the four cohorts is given in **Table S8**.

Supplemental figures

Figures

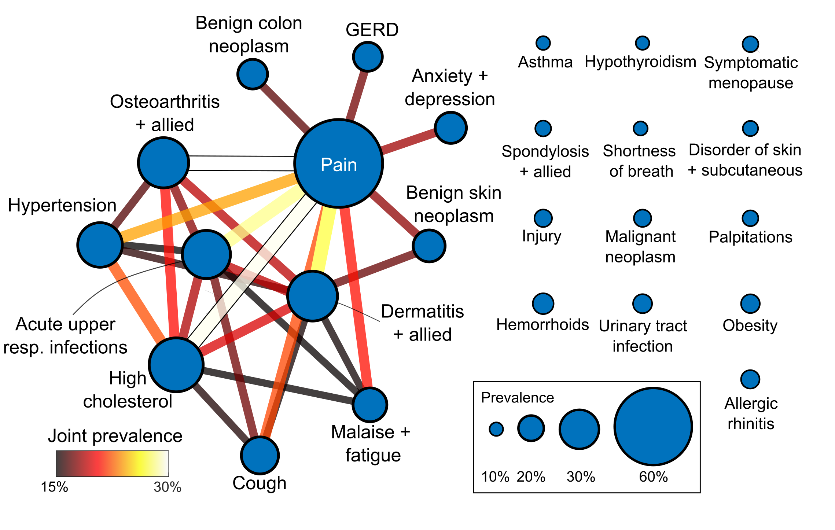

**Figure S1 | Common diagnoses in cohort A.** A network diagram of common diagnoses (based on PheCodes) in cohort A, during the study period (2002-2022). Edges reflect prevalence of patients with both diagnoses as a percentage of the total cohort with either (limited to edges above 15%).

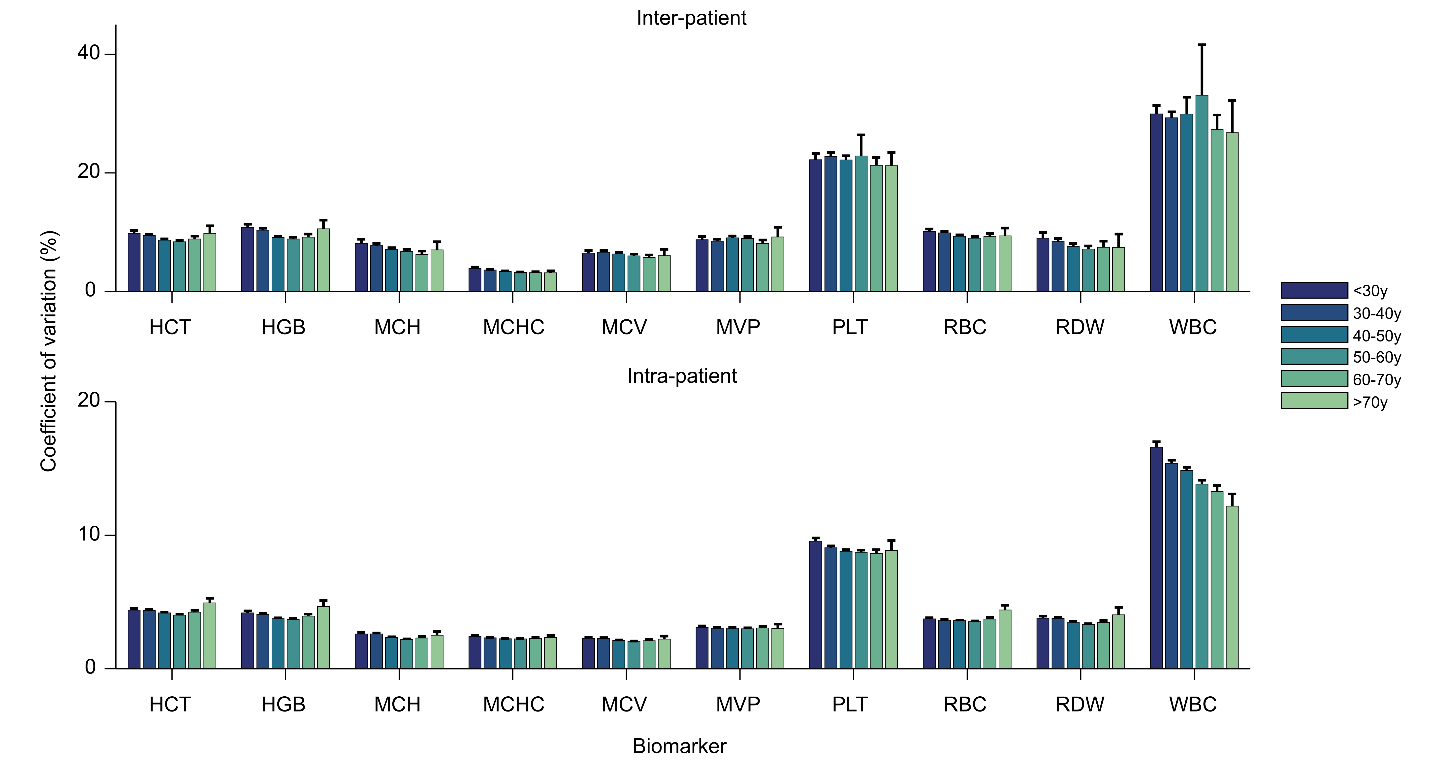

**Figure S2 | Long-term variation in blood count markers stratified by age.** Mean coefficients of variation for blood counts over a 20y period are given for Cohort A, stratified by their age at the study start point. Error bars represent 95% confidence intervals, generated via bootstrapping with 10,000 samples.

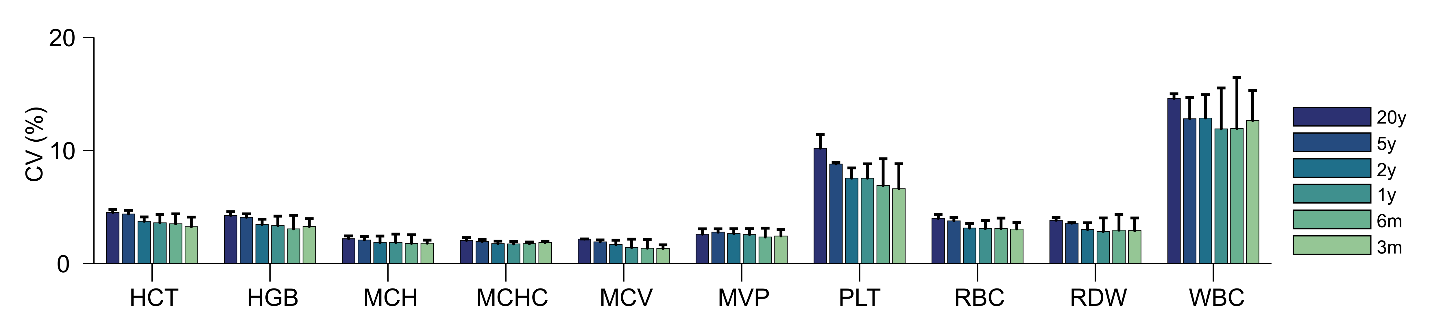

**Figure S3 | Intra-patient variation in blood count markers over various time periods.** Mean coefficients of variation for blood counts over long and short time periods. For each patient, the CV was calculated using the time period within the study with the highest number of isolated outpatient CBCs. Error bars represent 95% confidence intervals, generated via bootstrapping with 10,000 samples.

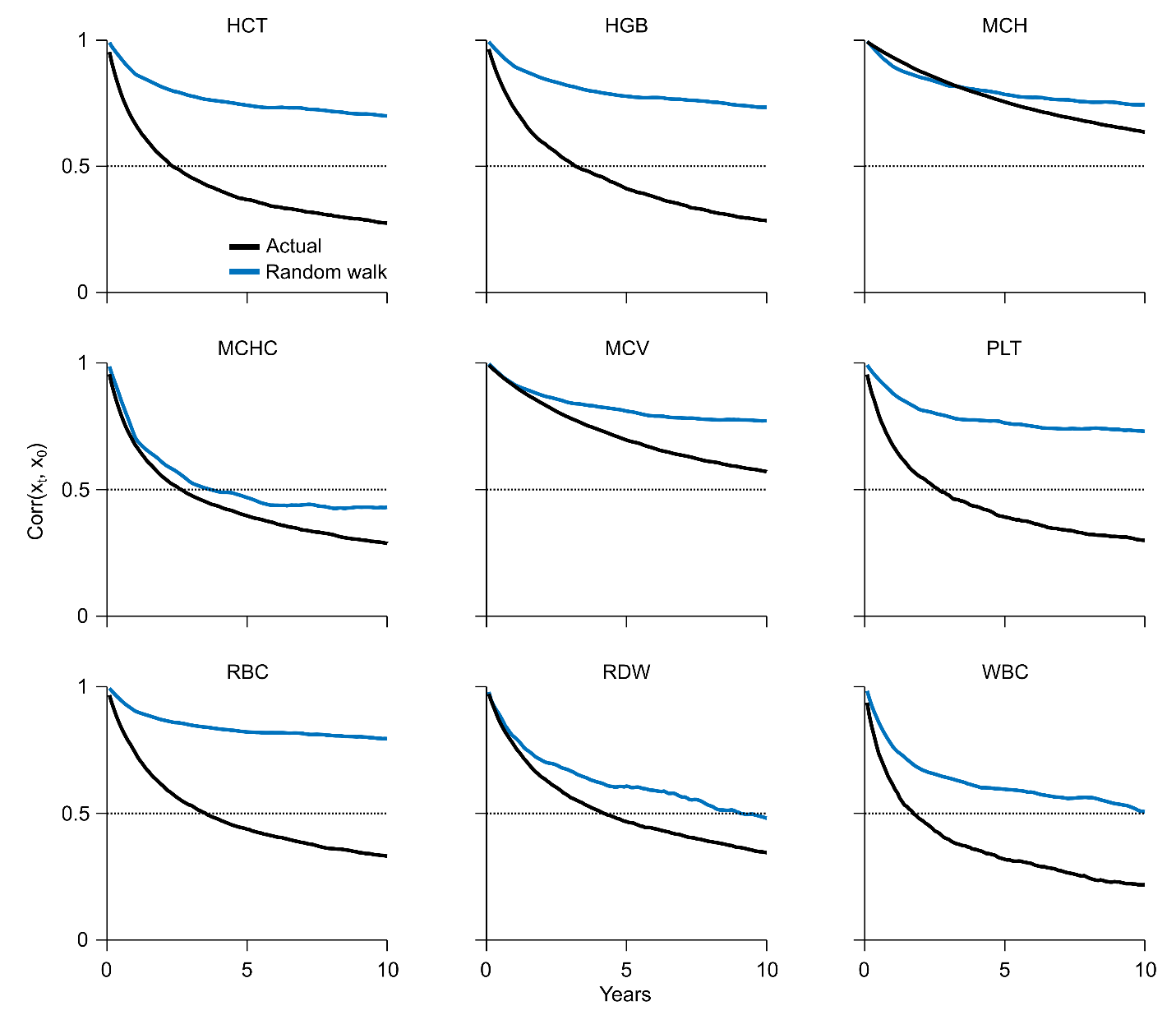

**Figure S4 | Correlations between markers over multiple years for cohort A and an equivalent random walk process.** Correlations between marker values in 2002 (year 0) and marker values at each year after for cohort A. The blue line represents the actual data, and the black line represents correlations for a random walk starting from the same values, with changes drawn each month from a normal distribution with mean 0 and standard deviation derived from the EFLM estimate of intra-patient marker CV. The setpoints exhibit substantially higher autocorrelation over long time periods than is seen for an equivalent random walk, consistent with the hypothesis that hematologic setpoints are actively regulated.

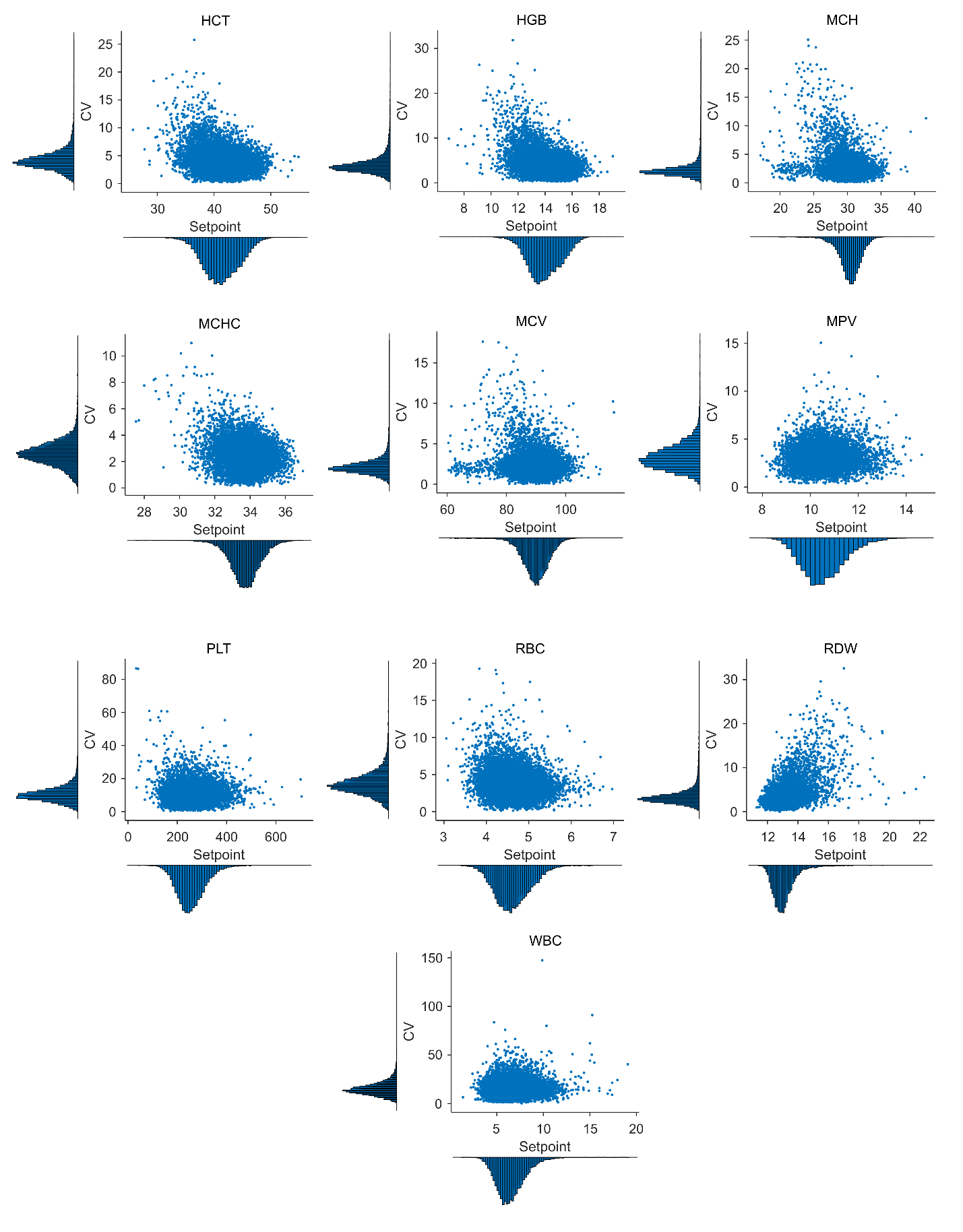

**Figure S5 | Association between setpoint and CV.** Plots of setpoint against CV are given for all patients in cohort A. Except for RDW, little correlation is seen between setpoint value and CV.

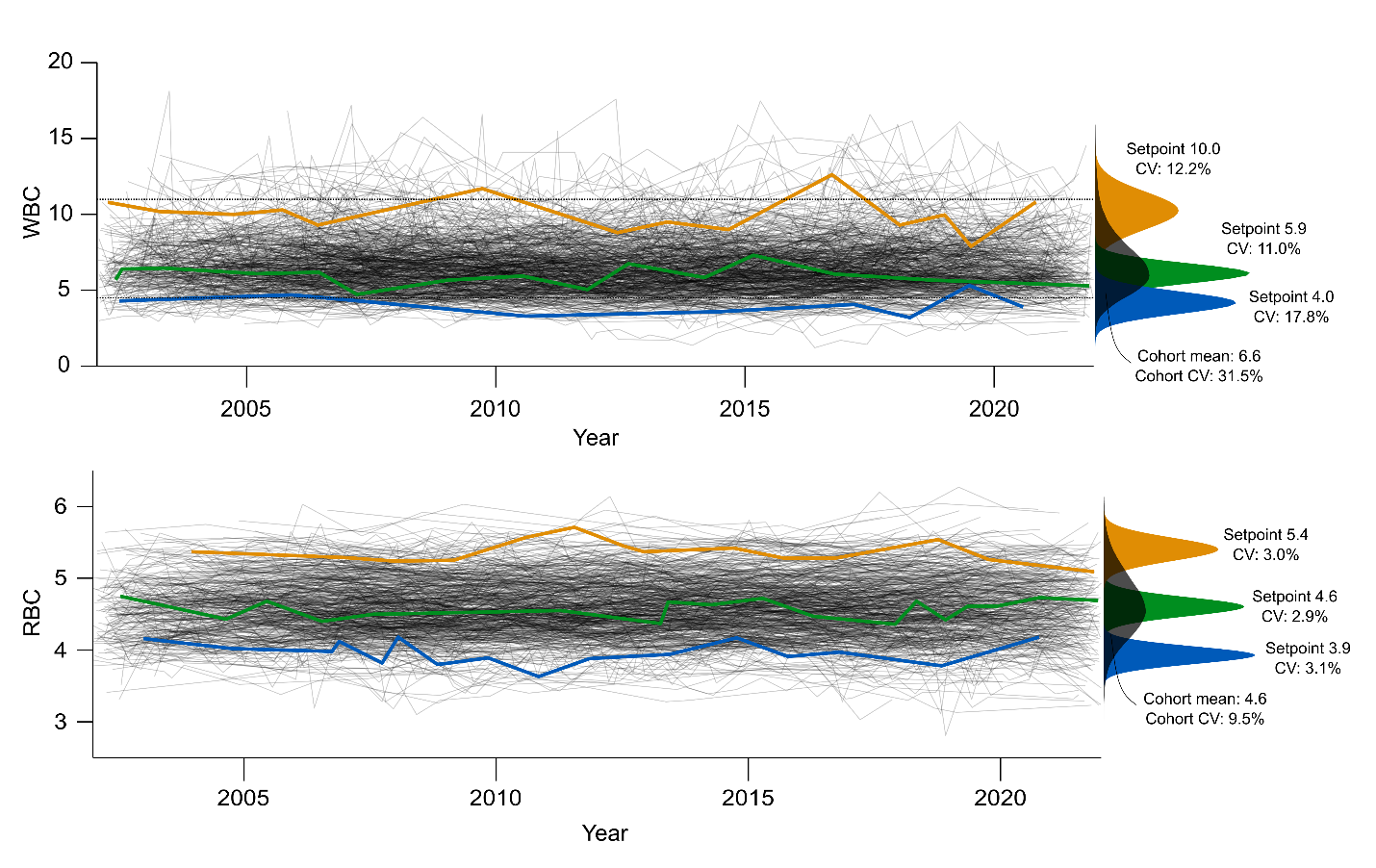

**Figure S6 | Outpatient trajectory plots for red and white cell counts.** Trajectory plots of all outpatient CBCs, similar to in **Fig 1e** for WBC and RBC. Each plot presents 500 randomly chosen patients in cohort A. Three patients have been highlighted, with setpoints in the lower, middle, or upper third of the cohort, and with all 3 having CVs between the 25-75^th^ percentiles for that marker.

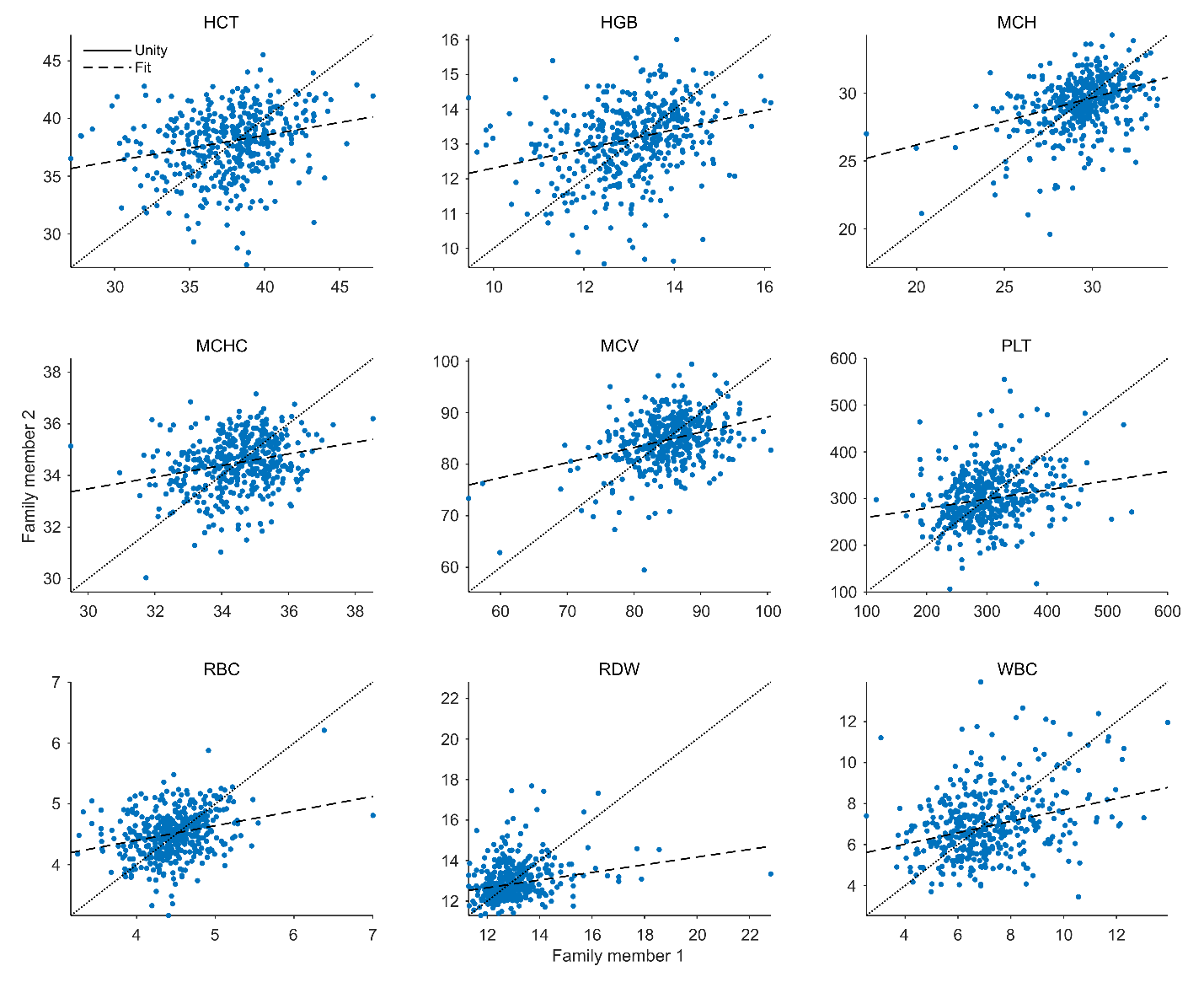

**Figure S7 | Correlation between blood count setpoints of first-degree relatives*.*** Associated correlation estimates are given in **Fig 4b**. Plotted values have been age- and gender-corrected via linear regression.

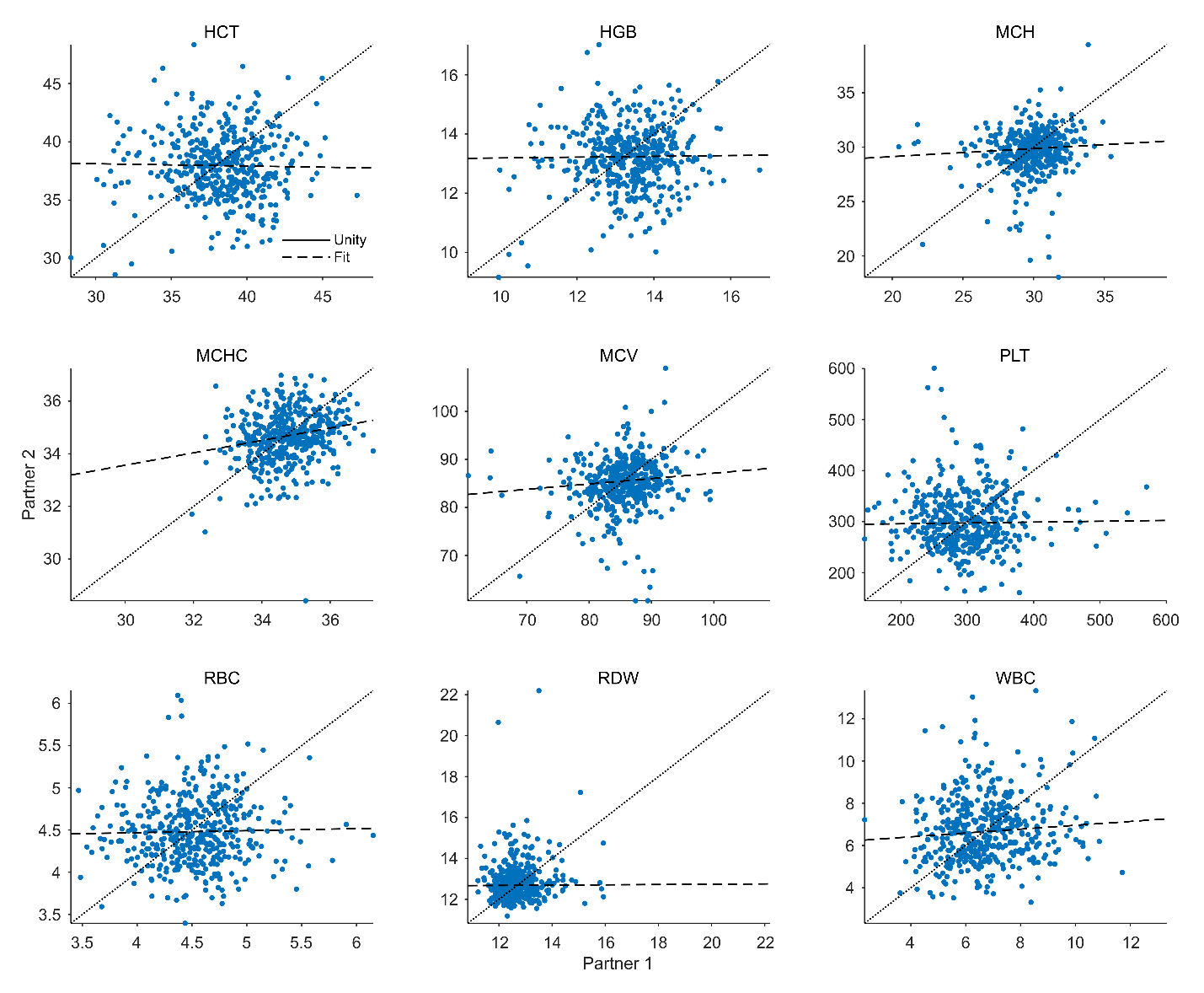

**Figure S8 | Correlation between blood count setpoints of spouses and life partners*.*** Associated correlation estimates are given in **Fig 4a**. Plotted values have been age- and gender-corrected via linear regression.

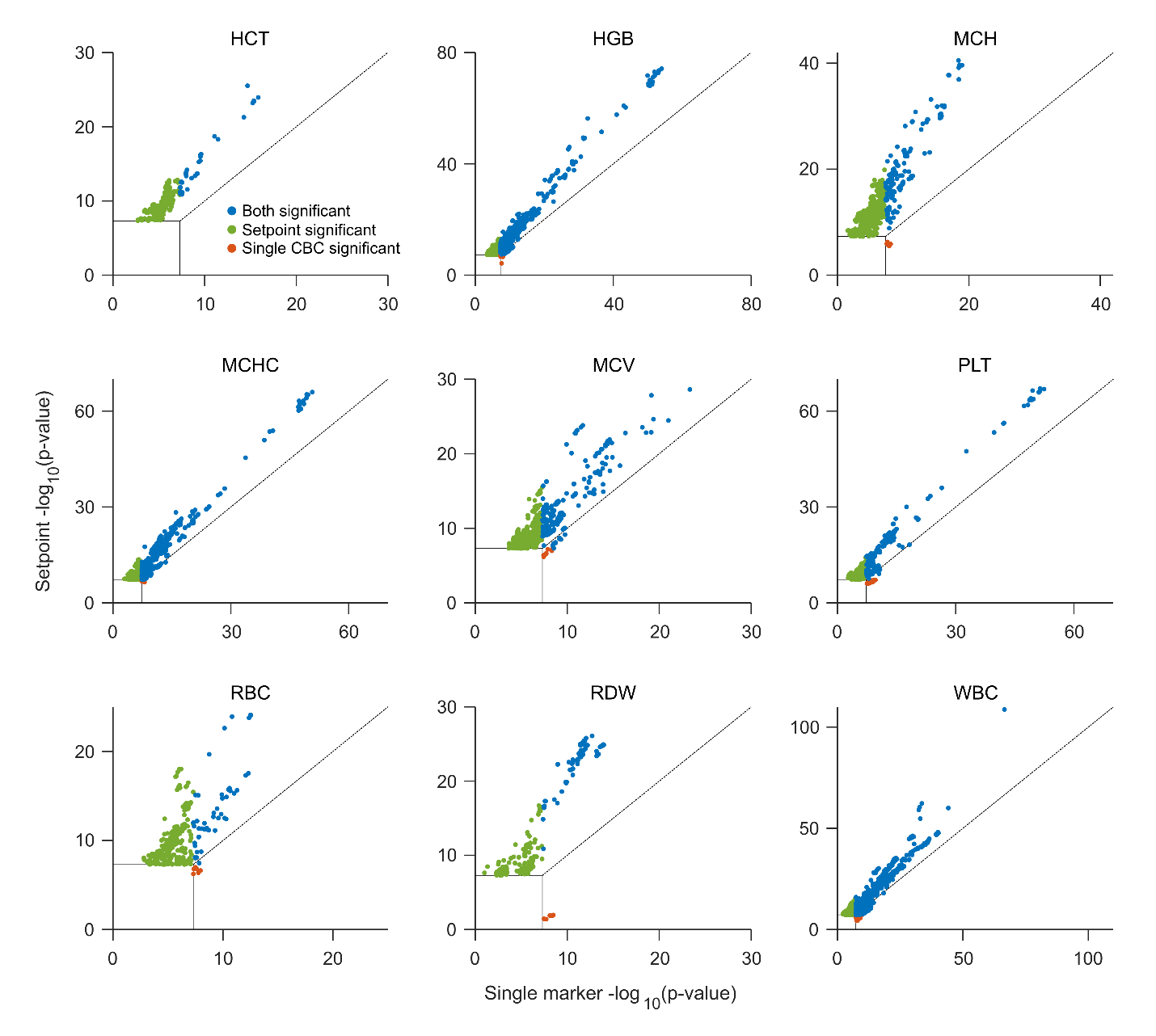

**Figure S9 | Relative significance of GWAS hits for CBC markers**. The relative significance of GWAS hits when using a marker setpoint comparative to a single isolated CBC chosen at random from the same study period. Dashed line represents unity, and highlighted colors reflect whether either or both markers achieved significance at p < 5e-8. Hits with both p-values below 5e-8 have been excluded for brevity.

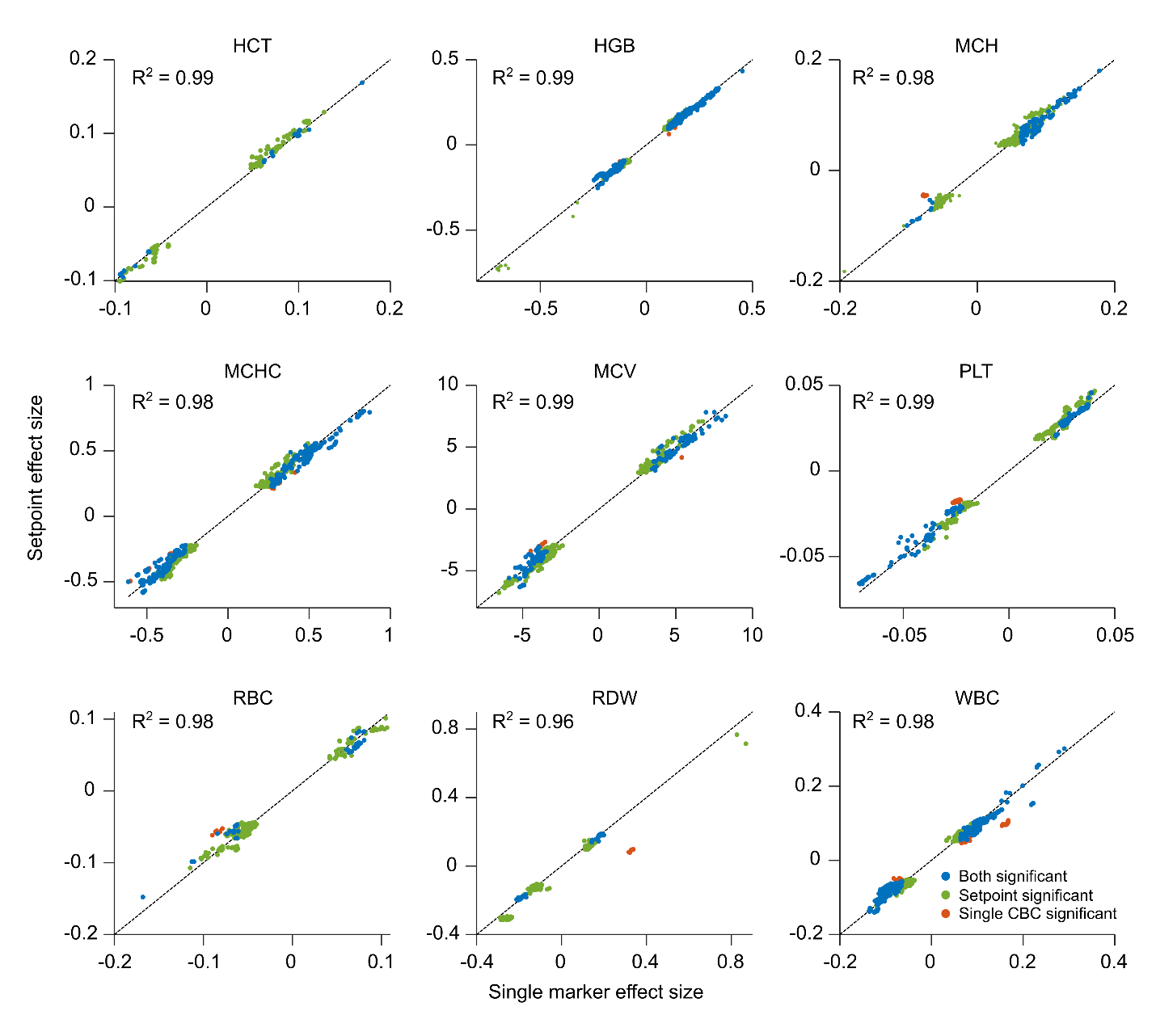

**Figure S10 | Comparison of effect sizes for significant GWAS hits**. Estimated effect sizes when using marker setpoints comparative to single isolated CBC chosen at random from the same study period. Markers are stratified by whether either or both p-values were significant at p < 5e-8. Dashed lines reflect unity.

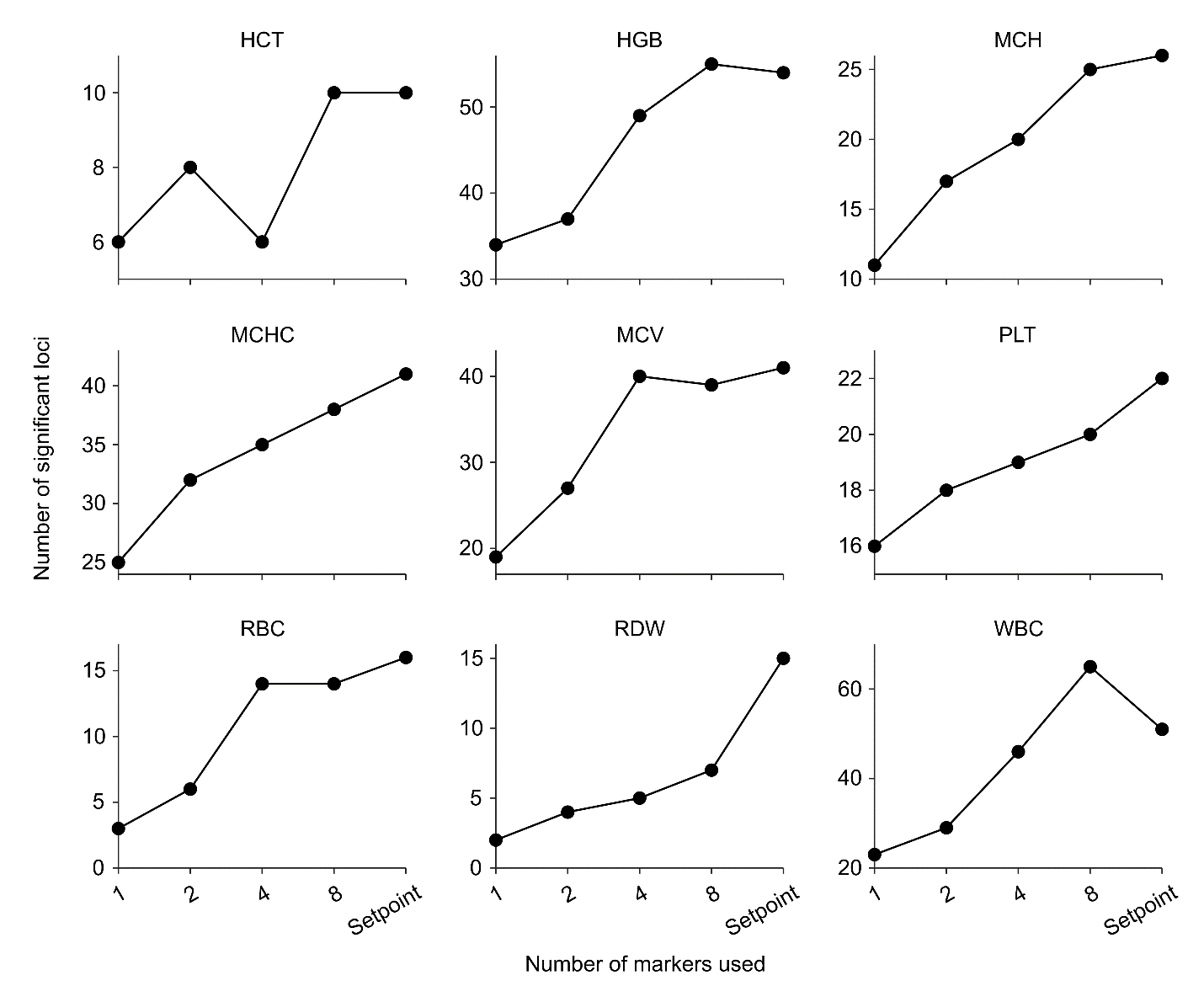

**Figure S11 | Number of significant loci identified when using averaged CBC values comparative to setpoints.** Results are from GWAS analyses following the same quality control measures as the primary analysis but limited to the patients with at least 8 isolated CBCs (N: 19,773). 1, 2, 4, and 8-point averages were taken from a randomly chosen subset of each patient’s isolated CBCs, but limited to the same set of 8 measurements, such that each higher point average contains all the data from the lower point average.

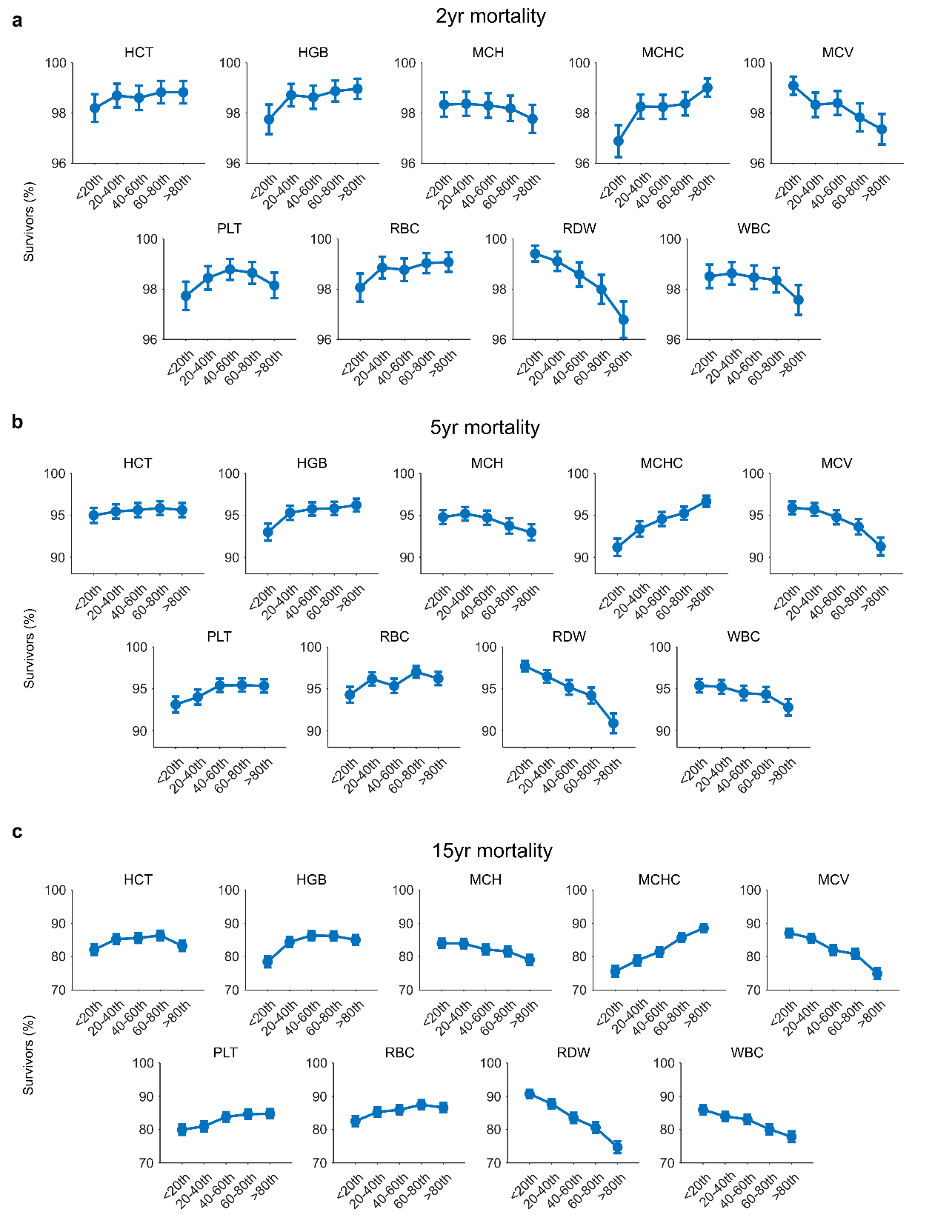

**Figure S12 | Setpoint-mortality associations over multiple time periods.** Patient mortality stratified by setpoint percentile in cohort B, over 2yrs (a), 5yrs (b), and 15yrs (c). Results are limited to setpoints within the MGH reference interval and use the percentile cut-offs defined in **Fig 5a**. Mortality associations are consistent across each of the time periods, with most markers showing strong monotonic associations.

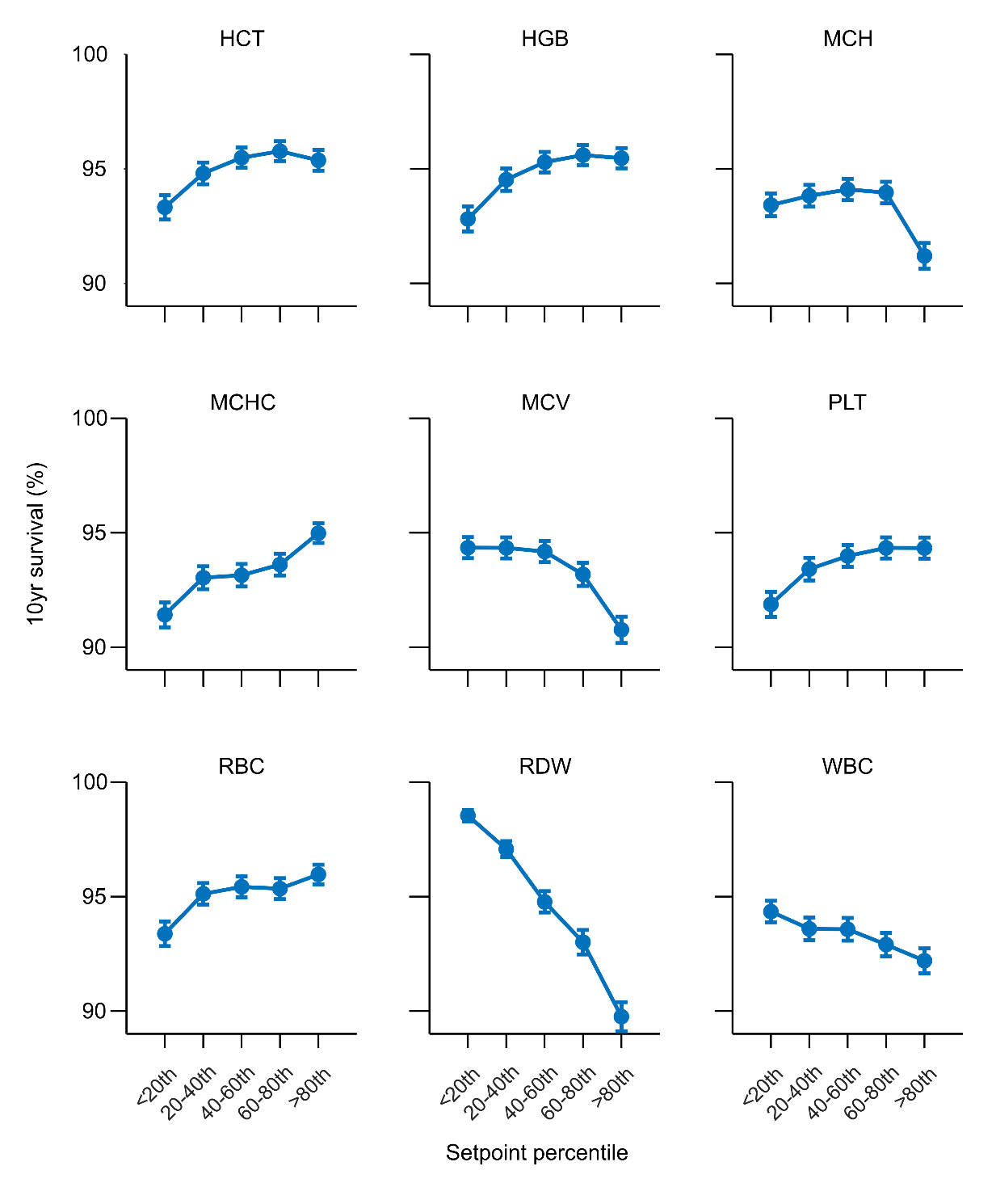

**Figure S13 | Validation of Fig 5b in an independent cohort.** 10yr survival rates stratified by setpoint percentile in a cohort of 50,423 individuals, limited to setpoints within the MGH reference interval. The cohort consists of all MGB patients with at least 5 isolated outpatient CBCs between 2006-2011, regardless of health status, but excluding any individuals in cohorts A-C. Survival rates are from Jan-01-2012 onwards. The curves are strikingly similar in shape to those in **Fig 5b**, with the same directional associations of setpoint values with general mortality. Note that overall mortality rates are lower in this figure, as this cohort is younger than cohort B (mean age 44.9yrs v 52.2yrs) and has a relatively smaller elderly population (10.0% > 65yrs old at study start, v 21.8% in cohort B).

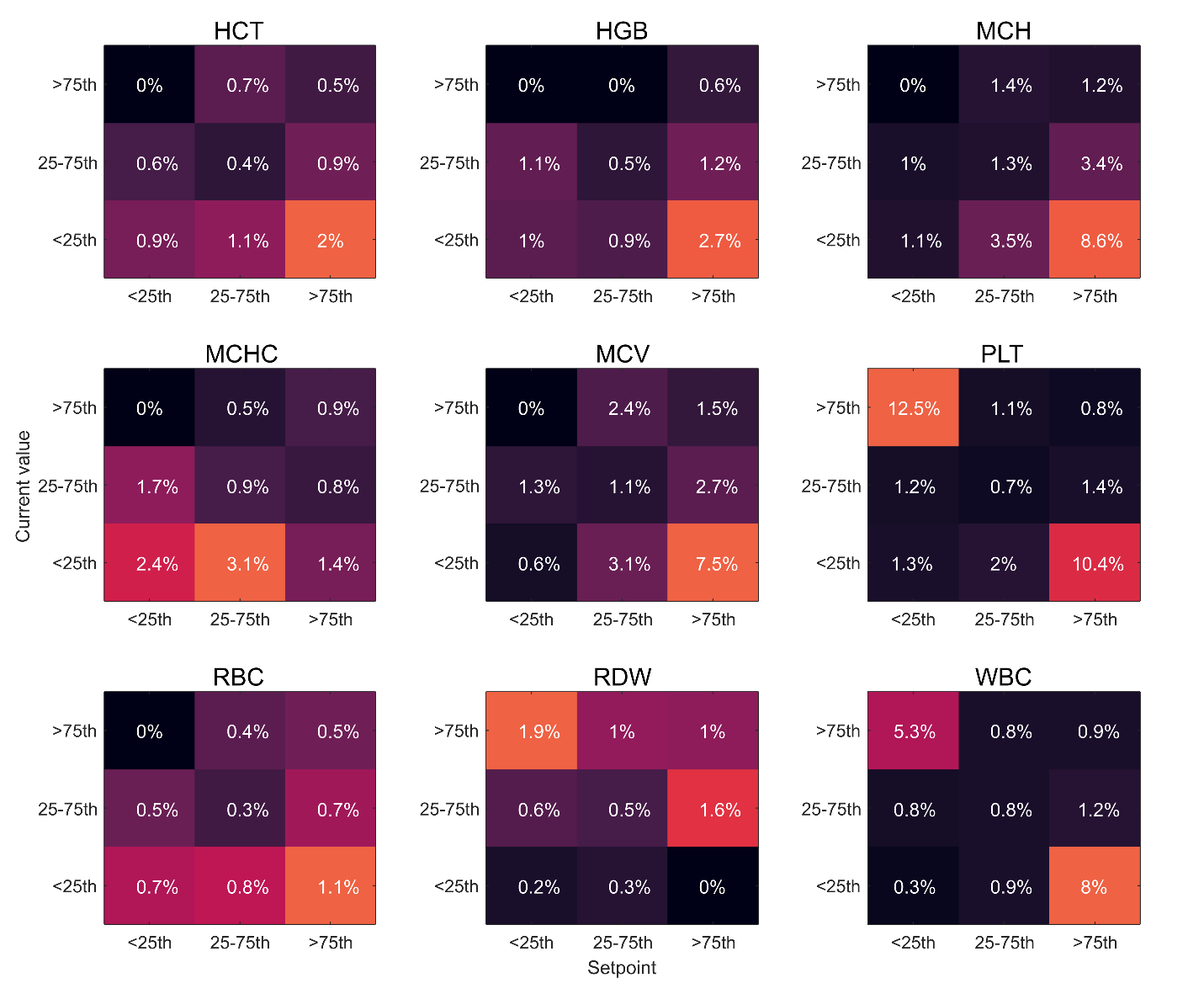

**Figure S14 | Mortality associations with setpoints and current marker values.** Plots of 1yr mortality, stratified by the marker setpoint (measured between 2002-2006) and the worst marker value in 2007. Plots are limited to marker values within the reference interval. For consistency across plots, cut-offs were set as the sex-specific 25^th^ and 75^th^ percentiles. 25^th^, 75^th^ percentiles (male/female) were: HCT: 42.4/37.8, 45.1/40.8; HGB: 14.4/12.8, 15.6/13.9; MCH: 30.0/29.4, 31.9/31.5; MCHC: 34.0/33.5, 35.1/34.5; MCV: 86.6/86.6, 92.1/92.3; PLT: 204/231, 271/309; RBC: 4.7/4.2, 5.1/4.6; RDW: 12.8/12.7, 13.6/13.6; WBC: 5.8/5.8, 7.8/7.9. Results for WBC may differ slightly to those in **Fig 5e**, as the cut-offs are slightly different, to allow for fair comparison to other markers in this figure. For most markers, high variability (either high setpoint and low value or vice versa) is associated with the highest mortality (HCT, HGB, MCH, MCV, PLT, RBC, RDW, WBC). As in **Fig 5e** many of the markers show strongly non-linear interactions, with the greatest risk being in patients with a high setpoint and low current value (HCT, HGB, MCH, MCV, RBC, WBC) or vice versa (PLT, RDW).

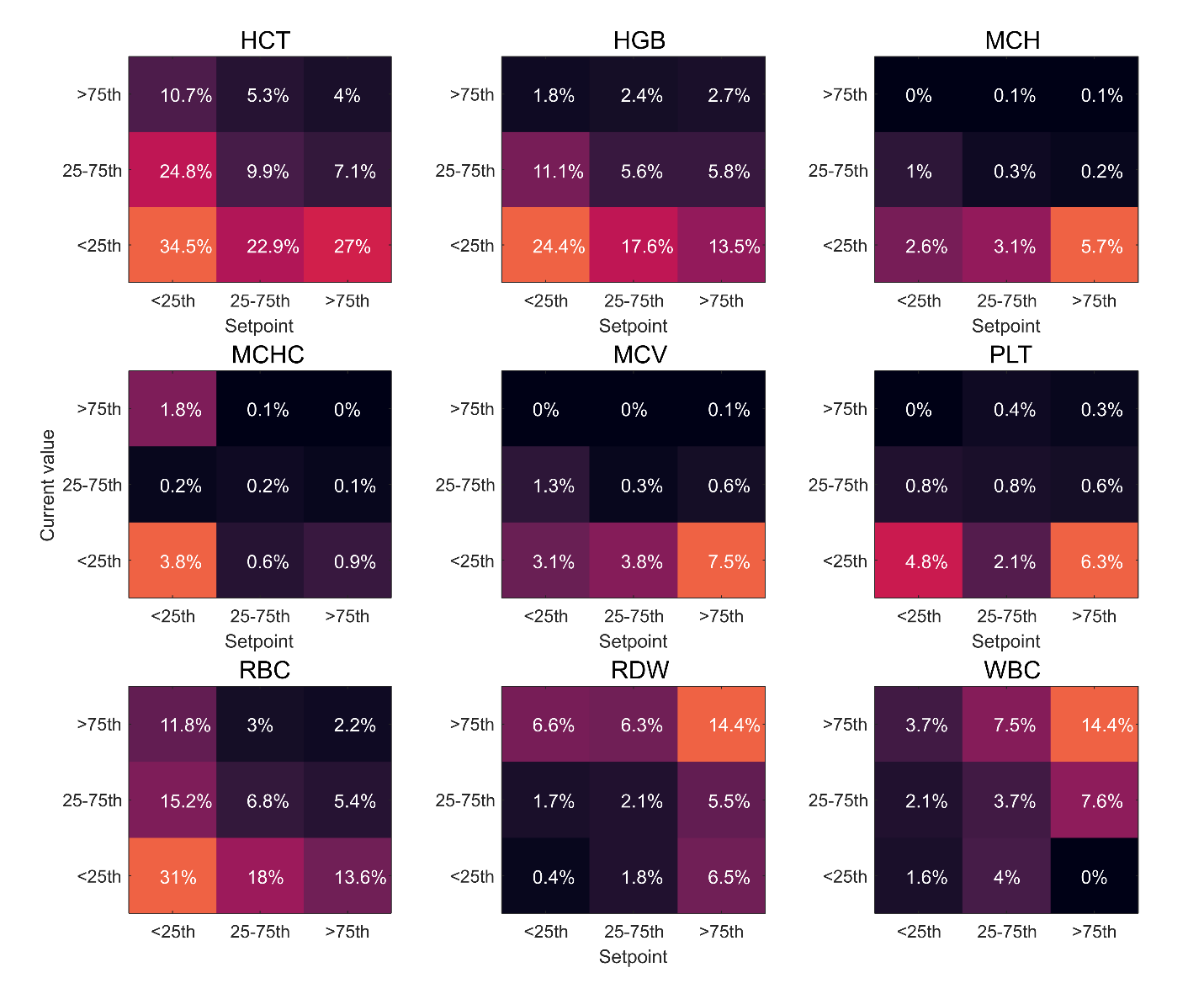

**Figure S15 | Lab abnormality associations with setpoints and current marker values.** Plots of 1yr risk of an abnormal lab test result, stratified by the marker setpoint (measured between 2002-2006) and the worst marker value in 2007. Lab abnormality was defined as an elevated (RDW, WBC) or depressed (HCT, HGB, MCH, MCHC, MCV, PLT, RBC) marker value, based on the MGH reference interval. Plots are limited to setpoint and marker values within the reference interval. For consistency across plots, cut-offs were set as the sex-specific 25^th^ and 75^th^ percentiles. 25^th^, 75^th^ percentiles (male/female) were: HCT: 42.4/37.8, 45.1/40.8; HGB: 14.4/12.8, 15.6/13.9; MCH: 30.0/29.4, 31.9/31.5; MCHC: 34.0/33.5, 35.1/34.5; MCV: 86.6/86.6, 92.1/92.3; PLT: 204/231, 271/309; RBC: 4.7/4.2, 5.1/4.6; RDW: 12.8/12.7, 13.6/13.6; WBC: 5.8/5.8, 7.8/7.9. Results for WBC may differ slightly to those in **Fig 5f**, as the cut-offs are slightly different, to allow for fair comparison to other markers in this figure. For some markers high discordance (a low setpoint and high value or vice versa) is associated with the largest risk (MCH, MCV, PLT) while for others, jointly elevated or depressed setpoints and current values are associated with the largest risk (HCT, HGB, MCHC, RBC, RDW, WBC). In most cases, risk is significantly non-linear, dependent on both setpoint and current marker values.

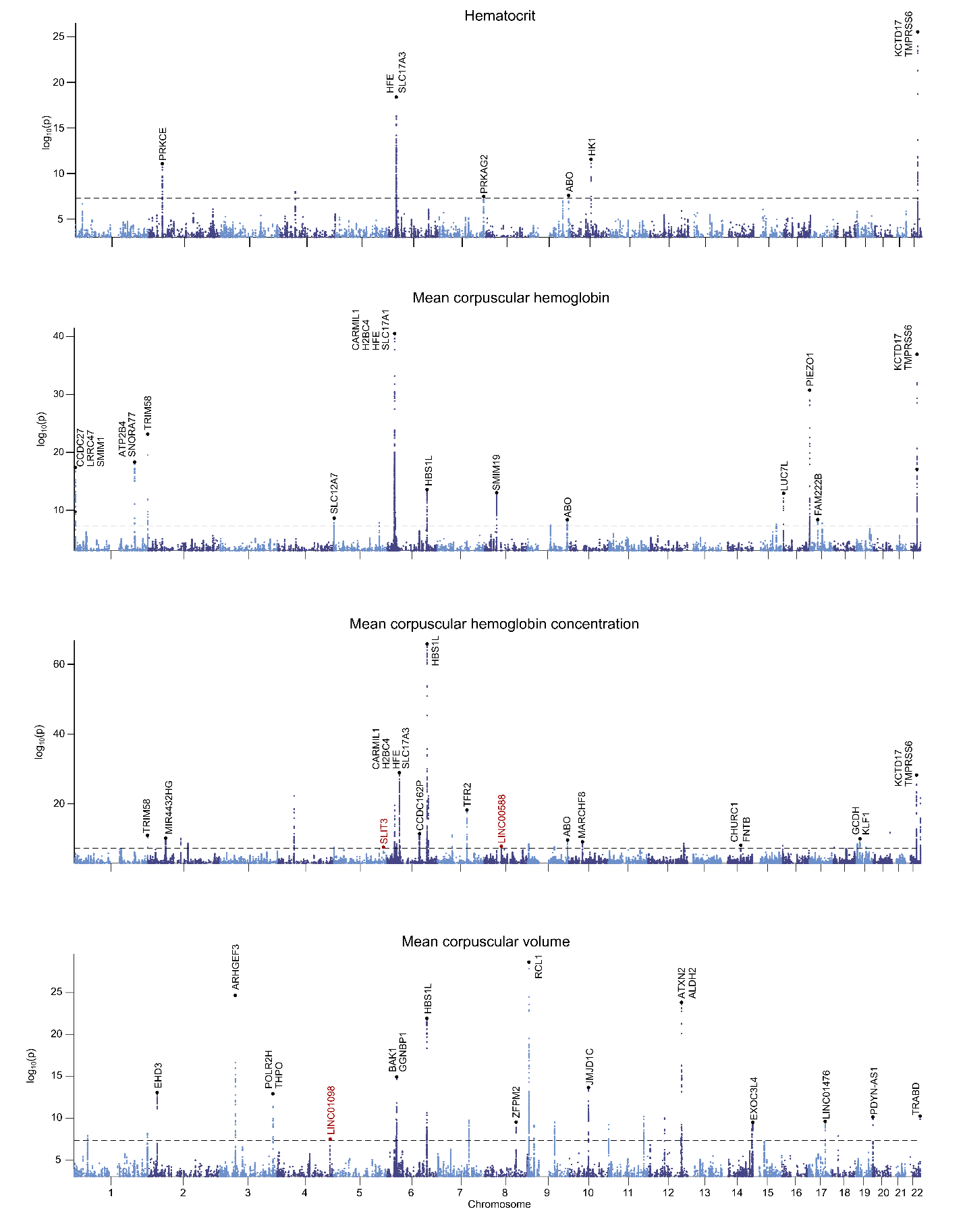

**Figure S16 | Manhattan plots for HCT, MCH, MCHC, and MCV.** Annotations correspond to nearby genes for the primary SNP in each association loci, for a collection of significant SNP clusters. Red annotations correspond to novel loci. A full list of significant hits, loci and nearby genes is given in **Tables S4-S6.**

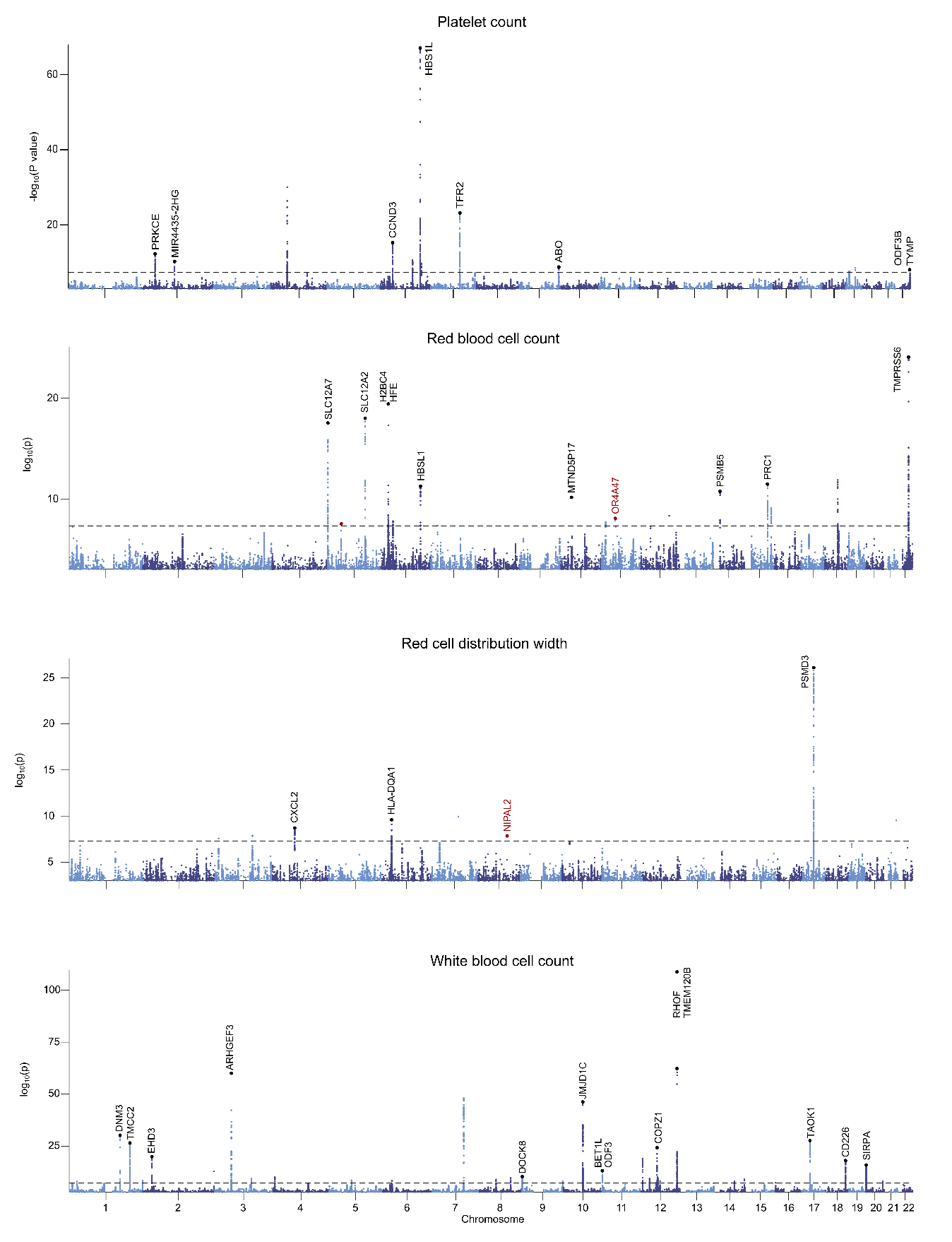

**Figure S17| Manhattan plots for PLT, RBC, RDW, and WBC.** Annotations correspond to nearby genes for the primary SNP in each association loci, for a collection of significant SNP clusters. Red annotations correspond to novel loci. A full list of significant hits, loci and nearby genes is given in **Tables S4-S6.**

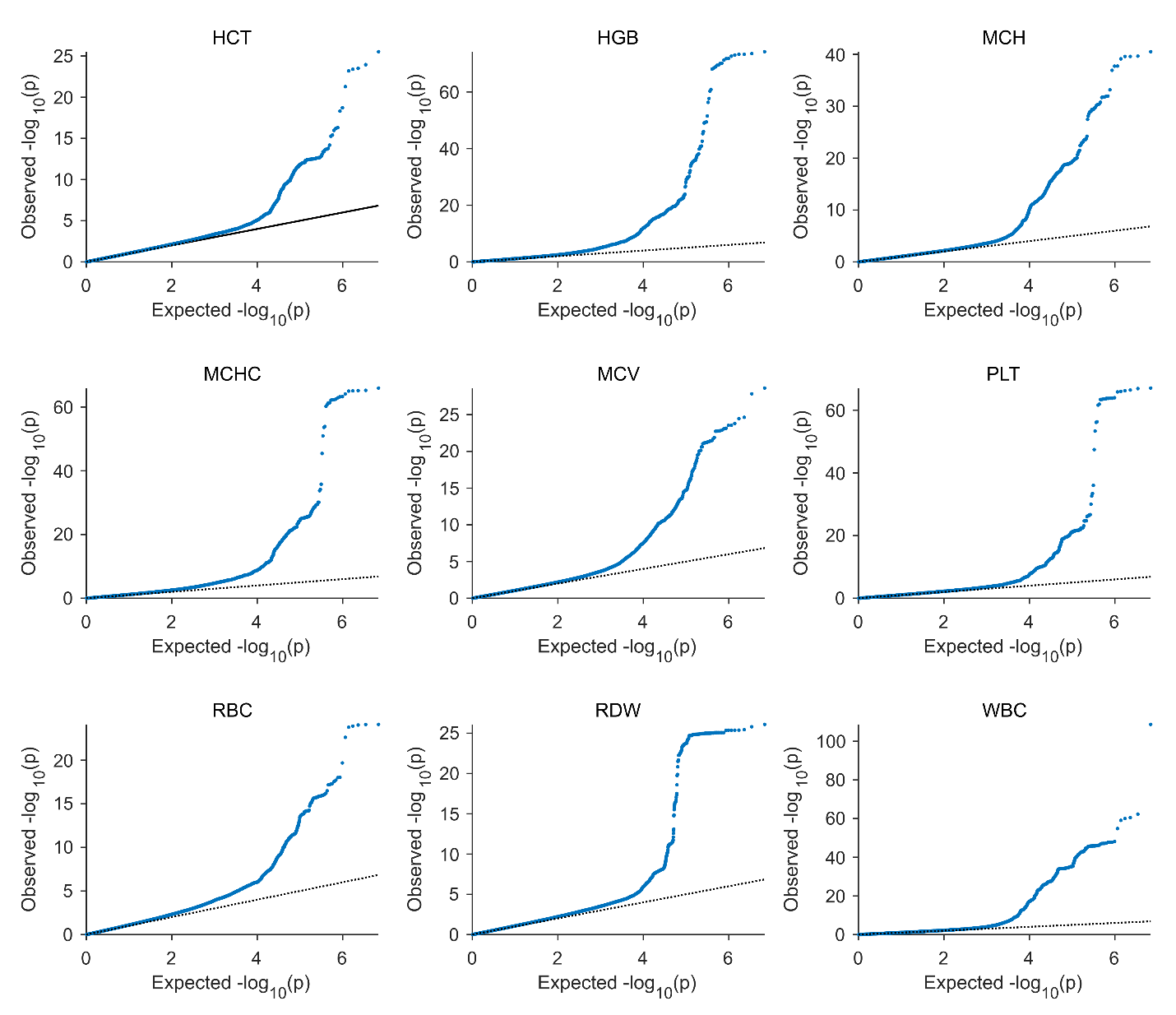

**Figure S18 | Quintile-quintile plots.** QQ-plots are given for p-values derived from each setpoint genome wide association study. In all cases none of the plots show significant evidence of confounding effects such as population stratification.

Supplemental tables

**Table S1 | Characteristics of primary study cohorts**

|  | Cohort A | Cohort B | Cohort C | Biobank cohort |
| --- | --- | --- | --- | --- |
| **Demographics** |  |  |  |  |
| N | 12407 | 14371 | 20062 | 25254 |
| Age – yrs | 43.3 (11.4) | 52.2 (15.3) | 40.9 (16.3) | 62.6 (15.5) |
| Sex - % male | 39.80% | 36.10% | 38.80% | 42% |
| Race - % white | 78.00% | 77.90% | 78.00% | 98% |
| Ethnicity - % Hispanic | 21.50% | 24.20% | 20.70% | 0.2% |
| **Setpoints** |  |  |  |  |
| # isolated CBCs | 14 (10-18) | 7 (6-9) | 8 (7-10) | 8 (13-22) |
| HCT | M: 43.8 (42.2-45.3) F: 39.8 (38.2-41.3) | M: 42.6 (40.5-44.4) F: 38.7 (36.7-40.4) | M: 43.1 (40.6-45.3) F: 39.7 (37.7-41.6) | M: 43 (41.1-44.8) F: 39.5 (37.9-41.1) |
| HGB | M: 15 (14.4-15.5) F: 13.4 (12.8-13.9) | M: 14.8 (13.9-15.4) F: 13.2 (12.4-13.8) | M: 14.4 (13.5-15.2) F: 13 (12.3-13.7) | M: 14.6 (13.8-15.2) F: 13.2 (12.6-13.7) |
| MCH | 30.4 (29.4-31.4) | 30.6 (29.4-31.7) | 30.2 (28.8-31.4) | 30.4 (29.4-31.4) |
| MCHC | 33.7 (33.2-34.2) | 34.2 (33.7-34.8) | 33 (32.3-33.6) | 33.5 (33-34.1) |
| MCV | 89.9 (87.1-92.7) | 89.3 (86.1-92.3) | 91.2 (87.9-94.5) | 90.6 (87.8-93.3) |
| PLT | 251.8 (218.5-290) | 256 (216.9-300.4) | 240.6 (200.6-285.6) | 247.8 (211.3-287.3) |
| RBC | M: 4.9 (4.7-5.1) F: 4.4 (4.2-4.6) | M: 4.8 (4.5-5) F: 4.4 (4.1-4.6) | M: 4.7 (4.4-5) F: 4.4 (4.1-4.6) | M: 4.8 (4.5-5) F: 4.4 (4.2-4.6) |
| RDW | 13 (12.6-13.4) | 13.2 (12.8-13.8) | 13.2 (12.6-13.9) | 13.2 (12.7-13.7) |
| WBC | 6.4 (5.5-7.4) | 6.7 (5.6-8) | 6.6 (5.5-7.9) | 6.9 (5.9-8.1) |
| **Outcomes** |  |  |  |  |
| 1yr mortality | 0.30% | 0.90% | 0.80% | 0% |

*Setpoint data is reported as median (25^th^-75^th^ percentiles), age is reported as mean (std). For cohorts A0-C age is at start of study, and for the biobank cohort age is on Jan-01-2022. 1yr mortality is from end of study. # of isolated outpatient CBCs is for during the study period for cohorts A-C, and over any time period for the biobank cohort. Note that the biobank cohort is limited to patients who passed the genomic quality control steps (see methods), including limiting to a European population. The 2% of non-white biobank subjects exclusively refers to patients who did not disclose race status in their medical record or listed multiple statuses.*

**Table S2 | Blood count reference intervals at Massachusetts General Hospital**

| Marker | Abbreviation | Units | Male | Female |
| --- | --- | --- | --- | --- |
| Hematocrit | HCT | % | 41-53 | 36-46 |
| Hemoglobin | HGB | g/dL | 13.5-17.5 | 12.0-16.0 |
| Mean corpuscular hemoglobin | MCH | pg | 26-34 | 26-34 |
| Mean corpuscular hemoglobin concentration | MCHC | g/dL | 31-37 | 31-37 |
| Mean corpuscular volume | MCV | fL | 80-100 | 80-100 |
| Mean platelet volume | MPV | fL | 8.4-12.0 | 8.4-12.0 |
| Platelet count | PLT | 10^3^/µL | 150-400 | 150-400 |
| Red blood cell count | RBC | 10^6^/µL | 4.5-5.9 | 4.0-5.2 |
| Red cell distribution width | RDW | % | 11.5-14.5 | 11.5-14.5 |
| White blood cell count | WBC | 10^3^/µL | 4.5-11.0 | 4.5-11.0 |

*Reference intervals are as of Jan-01-2023*

**Table S3 | Age and gender effects on marker values**

| Marker | Units | Median | Gender effect | Age effect | Age effect/median | Age effect/SD |
| --- | --- | --- | --- | --- | --- | --- |
| HCT | % | 41.1 | 3.94 | 0.023 | 0.1% | 0.5% |
| HGB | g/dL | 13.9 | 1.59 | -0.003 | 0.0% | -0.1% |
| MCH | pg | 30.5 | 0.49 | 0.009 | 0.0% | 0.4% |
| MCHC | g/dL | 33.7 | 0.64 | -0.025 | -0.1% | -1.0% |
| MCV | fL | 90 | -0.21 | 0.102 | 0.1% | 4.5% |
| MPV | fL | 10.5 | 0.02 | -0.002 | 0.0% | -0.1% |
| PLT | 10^3^/µL | 251 | -36.26 | -0.668 | -0.3% | -6.3% |
| RBC | 10^6^/µL | 4.57 | 0.45 | -0.003 | -0.1% | -0.1% |
| RDW | % | 13 | -0.19 | 0.007 | 0.1% | 0.2% |
| WBC | 10^3^/µL | 6.32 | -0.02 | -0.003 | -0.1% | 0.0% |

*Age and gender coefficients in a linear regression model over each set of markers in Cohort A. Age markers are per 1yr increase, gender markers are increase in male subjects comparative to females. Median marker values across the cohort are provided as a scale reference. Age effect is also presented as a percentage of the median and the average intra-patient standard deviation for the marker.*

**Table S4 | Sysmex setpoint summary characteristics**

| Marker | Description | Units | N | % Male | Age | Value |
| --- | --- | --- | --- | --- | --- | --- |
| ABASO | Absolute basophil count | 10^3^/µL | 4804 | 45.1% | 60.6 (16.4) | 0 (0-0.1) |
| AEOS | Absolute eosinophil count | 10^3^/µL | 4804 | 45.1% | 60.6 (16.4) | 0.1 (0.1-0.2) |
| ALYMPH | Absolute lymphocyte count | 10^3^/µL | 4804 | 45.1% | 60.6 (16.4) | 2.5 (2.1-2.9) |
| AMONO | Absolute monocyte count | 10^3^/µL | 4804 | 45.1% | 60.6 (16.4) | 0.5 (0.4-0.6) |
| ANEUT | Absolute neutrophil count | 10^3^/µL | 4804 | 45.1% | 60.6 (16.4) | 3.4 (2.6-4.5) |
| BASO | Basophil fraction | % | 4804 | 45.1% | 60.6 (16.4) | 0.7 (0.5-0.9) |
| EOS | Eosinophil fraction | % | 4804 | 45.1% | 60.6 (16.4) | 2.1 (1.4-3.2) |
| FRC | Fragmented red cell count | % | 92 | 43.5% | 65.3 (15.8) | 0.1 (0-1.8) |
| HFLC% | High fluorescence lymphocyte count | % | 3852 | 43.5% | 59.8 (16.8) | 0.1 (0-0.1) |
| IG# | Immature granulocyte count | 10^3^/µL | 4804 | 45.1% | 60.6 (16.4) | 0 (0-0) |
| IG% | Immature granulocyte fraction | % | 4804 | 45.1% | 60.6 (16.4) | 0.2 (0.2-0.3) |
| IPF | Immature platelet fraction | % | 292 | 57.9% | 65.6 (15.6) | 8.6 (6.3-13) |
| IRF | Immature reticulocyte fraction | % | 92 | 43.5% | 65.3 (15.8) | 9.3 (5.6-13.9) |
| LYMPH | Lymphocyte fraction | % | 4804 | 45.1% | 60.6 (16.4) | 38.4 (32.7-44.4) |
| MACRO | Macrocyte fraction | % | 8098 | 44.8% | 62.4 (16.2) | 3.8 (3.5-4.2) |
| MICRO | Microcyte fraction | % | 8098 | 44.8% | 62.4 (16.2) | 1.1 (0.6-2) |
| MONO | Monocyte fraction | % | 4804 | 45.1% | 60.6 (16.4) | 7.6 (6.2-9) |
| NEUT | Neutrophil fraction | % | 4804 | 45.1% | 60.6 (16.4) | 51.5 (45.1-57.4) |
| NEUT FSC | Median neutrophil forward scatter | ch | 4804 | 45.1% | 60.6 (16.4) | 90.7 (88.6-92.9) |
| NEUT SFL | Median neutrophil side fluorescence | ch | 4804 | 45.1% | 60.6 (16.4) | 46.7 (45.2-48.4) |
| NEUT SSC | Median neutrophil side scatter | ch | 4804 | 45.1% | 60.6 (16.4) | 153.3 (150.7-155.9) |
| NRBC% | Nucleated red cell fraction | % | 448 | 48.2% | 66.9 (15.3) | 0.1 (0-0.1) |
| PCT | Plateletcrit | % | 8044 | 44.7% | 62.4 (16.2) | 0.3 (0.3-0.4) |
| PDW | Platelet distribution width | fL | 8044 | 44.7% | 62.4 (16.2) | 12.4 (11.2-13.8) |
| RETIC | Reticulocyte count | % | 92 | 43.5% | 65.3 (15.8) | 1.5 (1.3-2) |
| RETHe | Mean reticulocyte hemoglobin | pg | 92 | 43.5% | 65.3 (15.8) | 33.3 (31.6-35.2) |
| RPI | Reticulocyte production index | % | 92 | 43.5% | 65.3 (15.8) | 1.3 (1-1.7) |

*N: number of patients in cohorts A-C with valid setpoints. Value: median (IQR) of setpoints. Age is mean(std) yrs. ch refers to measurements of normalized Sysmex channels, which use abstracted units. fL: femtolitres, pg: picograms*.

**Table S5 | Characteristics of the four prospective cohorts**

| **Cohort** | WBC | PLT | HCT | RDW |
| --- | --- | --- | --- | --- |
| **Demographics** |  |  |  |  |
| N | 20 | 20 | 20 | 18 |
| Age - yrs | 58.1 (11.0) | 51.7 (18.0) | 55.9 (11.9) | 65.6 (9.2) |
| Gender - % male | 50% | 50% | 50% | 44% |
| Race - % white | 90% | 100% | 90% | 89% |
| Ethnicity - % hispanic | 25% | 10% | 30% | 22% |
| **Setpoints** |  |  |  |  |
| HCT | 42.5 (39.8-45.5) | 42.3 (39.7-44.1) | 41.5 (39.6-42.3) | 42.6 (40.6-46.4) |
| HGB | 14 (13.1-15.1) | 13.9 (13-14.8) | 13.8 (13-14.1) | 14.0 (13.5-15.3) |
| MCH | 30.6 (29.2-30.8) | 30.3 (29.8-31.3) | 30.2 (29.9-31.1) | 30 (29.2-30.8) |
| MCHC | 33.3 (32.8-33.6) | 33.2 (32.5-33.8) | 33.3 (32.5-33.8) | 32.9 (32.6-33.4) |
| MCV | 91.4 (89.5-93.2) | 90.9 (88.7-93.7) | 90.2 (88.8-94.5) | 90.6 (89.7-93.5) |
| PLT | 240.8 (209.9-258.1) | 225.9 (203.6-270.6) | 260 (199.4-291.4) | 233.3 (195.8-265.1) |
| RBC | 4.6 (4.4-4.9) | 4.6 (4.4-4.9) | 4.7 (4.2-4.8) | 4.6 (4.5-5.1) |
| RDW | 13.1 (12.6-13.4) | 12.5 (12.2-12.9) | 13 (12.7-13.4) | 13 (12.8-13.9) |
| WBC | 6.4 (5.4-7.1) | 6.8 (5.9-7.3) | 6.4 (5.6-7.8) | 7.3 (6.4-8.1) |

*Legend: Data is presented as mean (std) for age, and median (25-75^th^ percentile) for setpoints.*

**Table S6 | Characteristics of the setpoint shift cohorts**

| **Cohort** | Splenectomy | Menopause | Pregnancy | Hypothyroidism | Liver disease |
| --- | --- | --- | --- | --- | --- |
| **Demographics** |  |  |  |  |  |
| N | 12 | 792 | 66 | 511 | 286 |
| Age - yrs | 50.4 (7.9) | 55.2 (8.5) | 49.1 (13) | 56.1 (11.5) | 53.8 (10.8) |
| Gender - % male | 25% | 0% | 0% | 27% | 48% |
| Race - % white | 100% | 78% | 77% | 84% | 77% |
| Ethnicity - % Hispanic | 25% | 21% | 32% | 18% | 20% |
| **Setpoints** |  |  |  |  |  |
| # CBCs | 19 (15.5-26.5) | 18 (15-24) | 20 (16-28) | 19 (16-25) | 20 (16-26) |
| HCT | 40.5 (37.3-42.6) | 38.4 (36.7-39.9) | 37.9 (36.4-39.3) | 39.3 (37.5-41.2) | 40.3 (38-42.8) |
| HGB | 14.2 (13.3-14.8) | 13.5 (12.8-14) | 13.1 (12.7-13.8) | 13.8 (13.2-14.6) | 14.2 (13.3-15.2) |
| MCH | 31.9 (30.8-32.5) | 29.8 (28.6-30.8) | 29.7 (28.7-30.8) | 30 (28.9-30.9) | 30.1 (28.9-31.1) |
| MCHC | 35 (34.9-35.4) | 34.9 (34.4-35.4) | 34.7 (34.3-35.2) | 35.1 (34.6-35.6) | 35.3 (34.7-35.8) |
| MCV | 90.4 (87.8-92.2) | 84.7 (81.4-87.2) | 85.1 (82.6-87.7) | 84.5 (81.6-87.1) | 84.4 (80.9-87.2) |
| MPV | 10.5 (9.8-12.1) | 10.6 (10-11.2) | 10.7 (9.9-11.2) | 10.6 (10-11.2) | 10.8 (10.2-11.3) |
| PLT | 355.1 (305-419.3) | 302.1 (268.4-335.8) | 291.6 (266.7-329) | 289.4 (254.9-329.9) | 281.4 (247.3-327.4) |
| RBC | 4.4 (4.2-4.7) | 4.6 (4.3-4.8) | 4.5 (4.3-4.7) | 4.7 (4.4-4.9) | 4.8 (4.6-5.1) |
| RDW | 13 (12.6-13.8) | 12.7 (12.3-13.2) | 12.7 (12.4-13.2) | 12.7 (12.3-13.2) | 12.7 (12.3-13.2) |
| WBC | 8.3 (6.8-10.4) | 6.5 (5.6-7.6) | 6.4 (5.8-7.5) | 6.6 (5.8-7.6) | 6.7 (5.8-7.8) |
| **Setpoint shift** |  |  |  |  |  |
| Marker | RDW | HCT | PLT | MCV | PLT |
| Pre-event | 12.8 (12.1-13.5) | 37.9 (36.1-39.4) | 309.4 (277.9-337.3) | 83.9 (81.2-86.8) | 291.1 (257.5-339.7) |
| Post-event | 13.4 (12.7-13.9) | 38.9 (37.1-40.7) | 284.4 (253.1-325.2) | 84.8 (82.1-87.8) | 275 (239.5-313.5) |
| Δ Setpoint | 0.49 (0.16-1.08) | 1.02 (-0.04-2.13) | -18.2 (-38.0-5.5) | 0.85 (-0.24-2.17) | -16.4 (-35-0.2) |
| p-value | 0.002 | <0.001 | <0.001 | <0.001 | <0.001 |

*Legend: Data for age is presented as mean(std), and for setpoints as median (25-75^th^ percentile). P-values correspond to a 1-sample t-test on the setpoint change from pre to post.*

**Table S7 | Literature estimates of blood count trait heritability**

|  | Study 1 | Study 2 | Study 3 | Study 4 | Study 5 | Average |
| --- | --- | --- | --- | --- | --- | --- |
| Study | Garner et al. ^1^ | Whitfield et al.^2^ | Remacha et al.^3^ | PT Williams^4^ | Cohen et al.^5^ |  |
| Study type | Twin | Twin | Family | Family | EHR |  |
| Study N | 775 | 412 | 935 | 3929 | 2.8 million |  |
| HGB | 0.37 | 0.59 | 0.32 | 0.375 | 0.425 | 0.416 |
| HCT | 0.3 | 0.65 | 0.36 | 0.375 | 0.387 | 0.4144 |
| MCH | 0.39 | 0.44 | 0.53 | 0.51 | 0.649 | 0.5038 |
| MCHC | 0 | 0.14 | 0.54 | 0.235 | 0.495 | 0.282 |
| MCV | 0.2 | 0.47 | 0.55 | 0.475 | 0.642 | 0.4674 |
| MPV | NA | 0.88 | NA | 0.69 | 0.522 | 0.697333 |
| PLT | 0.57 | 0.86 | NA | 0.37 | 0.607 | 0.60175 |
| RBC | 0.42 | 0.75 | 0.44 | 0.475 | 0.585 | 0.534 |
| RDW | NA | NA | 0.49 | 0.08 | 0.419 | 0.329667 |
| WBC | 0.62 | 0.58 | NA | 0.41 | 0.49 | 0.525 |

EHR: Study using large-scale analysis of electronic health records. NA: study didn’t report estimate

**Table** **S8, S9** and **S10** are included in excel file **Supplemental File 2**.
