## Supplemental File 3 for "Hematologic setpoints are a stable and patient-specific deep phenotype": CalculateSetpoint.m

```
function [setpoint, setpoint_cv] = CalculateSetpoint(x, y)
% FUNCTION [setpoint, setpoint_cv] = CalculateSetpoint(x, y) estimates the
% setpoint of a time series defined by dates X, and values y. 
% 
% INPUTS:
% x: A vector of sampling times. If the vector is a datetime, it will be
% converted to a numerical array. 
% y: A vector of marker values, of same length as x. 

% OUTPUTS: 
% setpoint: The estimated setpoint for the time series
% setpoint_cv: The estimated coefficient of variation for the setpoint
% returned as a decimal percentage
%
% This code was written as part of a study titled: 
% Hematologic setpoints are a stable and patient-specific deep phenotype
% Written by Dr Brody H Foy (research fellow, Higgins Lab, Harvard Medical 
% School & Massachusetts General Hospital), in 2023. 
% contact:, www.brodyfoy.com/
%
% If using this code for research purposes, please cite the publication: 
% BH Foy et al., Hematologic setpoints are a stable and patient-specific deep phenotype
%
% This code is provided with a creative commons license, and is free to
% use and modify in any way. The authors make no claims of performance or
% accuracy for any commercial or non-commercial contexts. 

%% Define basic parameters
min_marker_gap = 10; %Limit to markers at least this many (presumed units) days
% apart
min_tests = 5; %The minimum number of markers needed to calculate the setpoint (default 5)


%% Check for viability
x = x(:); y = y(:); %Convert to column vectors
idx = ~isnan(y); % Remove any NaN data and sort into order of x
x = x(idx); y = y(idx); 

[x, unique_idx] = unique(x);
y = y(unique_idx); 

[x, sort_idx] = sort(x); 
y = y(sort_idx);

% Case from datetime to double if necessary
if isdatetime(x)
    x = days(x - x(1));
end

%% Limit to markers within the desired gap
nearest_marker = NaN(length(x),1); 
for i = 1:length(x)
    nearest_marker(i) = min(abs(x(i) - setdiff(x, x(i)))); 
end
x = x(nearest_marker > min_marker_gap); 
y = y(nearest_marker > min_marker_gap); 

%% If not enough data passes the filters, throw error
if length(x) < min_tests
    error(['Less than ', num2str(min_tests), ' data points are valid']); 
end

%% Calculate setpoint
mdl = cell(3,1); %Holds mixture components
aic = NaN(3,1); %Holds AIC values for each model
for k = 1:3
    try
        mdl{k} = fitgmdist(y, k, 'Options', statset('MaxIter', 300), 'RegularizationValue', 0.001); 
        aic(k) = mdl{k}.AIC; 
    end
end

%% Select the best model
[~, min_idx] = min(aic); 
mdl = mdl{min_idx}; 
mdl_prop = mdl.ComponentProportion; %Size of biggest component

if (min_idx == 2 && max(mdl_prop) > 0.7) || (min_idx == 3 && max(mdl_prop) > 0.45)
    [~, max_idx] = max(mdl_prop); %Choose largest component
    setpoint = mdl.mu(max_idx); 
    setpoint_cv = sqrt(mdl.Sigma(max_idx))/(mdl.mu(max_idx)); 
else 
    % If multi-component model was invalid, use the data mean
    setpoint = mean(y);
    setpoint_cv = std(y)/setpoint; 
end
```
